# Comparative analysis unveils novel changes in serum metabolites and metabolomic networks of retinopathy of prematurity infants

**DOI:** 10.1101/2021.04.22.21255917

**Authors:** Yuhang Yang, Qian Yang, Yinsheng Zhang, Chaohui Lian, Honghui He, Jian Zeng, Guoming Zhang

## Abstract

**Background:** Advances in mass spectrometry are providing new insights into the role of metabolomics in the aetiology of many diseases. Studies in retinopathy of prematurity (ROP), for instance, overlooked the role of metabolic alterations in disease development. Here, we employed comprehensive metabolic profiling and gold-standard metabolic analysis to explore major metabolites and metabolic pathways significantly affected in early stages of pathogenesis toward ROP.

**Methods:** This is a multicentre, retrospective case-control study. We collected serums from 57 ROP cases and 57 strictly baseline matched non-ROP controls. Non-targeted ultrahigh performance liquid chromatography-tandem mass spectroscopy (UPLC-MS/MS) from Metabolon, Inc. was used to detect the metabolites in serum samples. Machine learning was used to unravel most affected metabolites and pathways in ROP development.

**Results:** Compared to non-ROP controls, we found a significant metabolic perturbation in the ROP serums, featured with an increase in lipid, nucleotide, carbohydrate metabolites and a lower level of peptides. Machine leaning helped to distinguish a cluster of metabolic pathways (glycometabolism, redox homeostasis, lipid metabolism and arginine pathway) that were strongly related to the development of ROP. In addition, we found that the severity of ROP was related to the level of creatinine and ribitol.

**Conclusion:** In the current study, our results suggested a strong link between metabolic profiling and retinal neovascularization during ROP pathogenesis. These findings provided an insight into identifying novel metabolic biomarkers for ROP diagnosis and prevention.

## Introduction

Retinopathy of prematurity (ROP) is characterized by the presence of retinal ischemia and retinal neovascularization which may lead to a retinal detachment which was first reported by Terry since the forties of the last century. It is a potential blind retinopathy disease that occurs commonly in preterm or preterm very low birth weight (VLBW) infants^[22, 26]^. Despite of improvement of neonatal intensive care and increased survival rates of VLBW infants in low- and middle-income countries^[4, 19]^, the incidence and severity of ROP maintained at a high level^[10, 46]^. A recent epidemiological study reported that, worldwide, approximately 100,000 children suffering from blindness are due to ROP^[26, 31]^.

There are some limitations of current screening and treatment strategies of ROP. Clinically diagnosis of ROP relies on indirect ophthalmoscopy and colour fundus images following the International Classification of ROP (ICROP)^[37]^, yet these two methods merely provide information about pathological change in the eye. Current therapeutic approaches to ROP include retinal laser photocoagulation and intravitreal injection of anti-vascular endothelial growth factor (VEGF) agents. However, risks have been identified on damaging the iris, lens and choroidal rupture by laser photocoagulation, which may also lead to constricted visual field and refractive errors in the future^[52]^. Injection of anti-VEGF agent to premature infants has raised concerns about infection, suppression on systemic normal vascular development and even inhibiting the growth of central nervous system^[21, 43]^. Furthermore, current treatments only aim at those who have already developed ROP, at which stage the damage in the eye is irreversible. Therefore, it would of urgent and great need to develop a preventative tactic to identify high-risk groups at an earlier stage and prevent disease progression effectively.

Metabolomics aims at the holistic measurement of a considerable number of small molecules (metabolites) from a biological sample. It is a rapidly evolving field in biochemical research and is regarded as a complementary technology to other “omics” techniques^[50]^. To the best of our knowledge, this is the first study performing metabolomic analysis on serums from ROP and control infants and comprehensively compare alteration in the metabolic profiles between groups; we aim to provide an insight into early diagnosis and treatment by identifying specific biomarkers and relevant pathways.

## Methods

### 2.1. Study Design

This is a multicentre case-control study with data collected between April 2018 and October 2019 from neonatal intensive care units (NICU) of 22 hospitals in Shenzhen, China. This study, adhered to the tenets of the Declaration of Helsinki, has been reviewed and approved by the medical ethics committee of all hospitals and has been registered at ClinicalTrials.gov (ChiCTR1900020677). Signed enrolment consent forms were obtained from all families. Infants were enrolled if they were born preterm (between 20 weeks to 37 weeks from gestation). A total of 114 patients fulfilled the inclusion criteria. The presence of ROP was determined by specialized ophthalmologists from Shenzhen Eye Hospital. The classification criteria were as follows: (1) ROP group: Severe ROP that required treatment (aggressive posterior retinopathy of prematurity (AP-ROP), zone I ROP requiring treatment or zone II ROP that required treatment (posterior pole); (2) control group: infants in control group had no ROP until the age of 40 PMA week. This control group was strictly matched with the ROP group for birth weight (BW) no greater than 200g difference and gestational age (GA) and postmenstrual age (PMA) of no more than 2-week difference. Participants will be excluded from the study if (1) preterm infants who had received any specific ocular treatment for ROP (including photocoagulation, vitrectomy, and intravitreal injections); (2) requested by participants’ families; (3) participants were diagnosed with congenital metabolomic diseases; (4) occurrence of severe complications, including but not limited to, sepsis, necrotizing enterocolitis, neonatal respiratory distress syndrome, and severe metabolic disorder; (5) the mother of the participant had a history of medication and/or serious disease, such as gonorrhoea, syphilis, or AIDS during pregnancy; (6) control group has ROP occurrence in subsequent follow-up visits.

### 2.2. Diagnosis of ROP

The diagnostic and therapeutic criteria of ROP follow the international classification^[1, 37]^ and the screening guidelines for retinopathy of prematurity in China (2014)^[17]^. All infants were screened using a binocular indirect ophthalmoscope (Heine, Bavaria, Germany) and RetCam (Natus Retcam3, California, United States) until the end of the follow-up. Every infant was examined by two experienced retina specialists independently and the eligibility of participation was confirmed by both the specialists. In our study, the severity of ROP was graded according to the guideline-based on the following criteria.(**Supplementary Table 1**)

### 2.3. Metabolic profiling

Sample preparation was carried out at Metabolon, Inc. as follows^[14]^: recovery standards were added prior to the first step in the extraction process for quality control purposes. To remove protein, dissociate small molecules bound to protein or trapped in the precipitated protein matrix, and to recover chemically diverse metabolites, proteins were precipitated with methanol under vigorous shaking for 2 min (Glen Mills Genogrinder 2000) followed by centrifugation. The resulting extract was divided into five fractions: two for analysis by two separate reverse phases (RP)/UPLC-MS/MS methods with positive ion mode electrospray ionization (ESI), one for analysis by RP/UPLC-MS/MS with negative ion mode ESI, one for analysis by HILIC/UPLC-MS/MS with negative ion mode ESI, and one sample was reserved for backup. Samples were placed briefly on a TurboVap® (Zymark) to remove the organic solvent. The sample extracts were stored overnight under nitrogen before preparation for analysis.

Three types of controls were analysed in concert with the experimental samples: 1) a pooled matrix sample generated by taking a small volume of each experimental sample served as technical replicate throughout the dataset; 2) extracted water samples served as process blanks; and 3) a cocktail of quality control standards that were carefully chosen not to interfere with the measurement of endogenous compounds were spiked into every analysed sample, allowed instrument performance monitoring and aided chromatographic alignment. Instrument variability was determined by calculating the median relative standard deviation (RSD) for the standards that were added to each sample before injection into the mass spectrometers (median RSD = 6%). Overall process variability was determined by calculating the median RSD for all endogenous metabolites (i.e., non-instrument standards) present in 100% of the pooled human plasma samples (median RSD = 8%). Experimental samples and controls were randomized across the platform run.

### 2.4. Ultrahigh Performance Liquid Chromatography-Tandem Mass Spectroscopy(UPLC-MS/MS)

The UPLC-MS/MS platform utilized a Waters ACQUITY UPLC and a Thermo Scientific Q-Exactive high resolution/accurate mass spectrometer interfaced with a heated electrospray ionization (HESI-II) source and Orbitrap mass analyser operated at 35,000 mass resolution. The sample extract was dried then reconstituted in solvents compatible to each of the four methods. Each reconstitution solvent contained a series of standards at fixed concentrations to ensure injection and chromatographic consistency. One aliquot was analysed using acidic positive ion conditions, chromatographically optimized for more hydrophilic compounds. In this method, the extract was gradient eluted from a C18 column (Waters UPLC BEH C18-2.1×100 mm, 1.7 µm) using water and methanol, containing 0.05% perfluoropentanoic acid (PFPA) and 0.1% formic acid (FA). Another aliquot was also analysed using acidic positive ion conditions and the extract was gradient eluted from the same aforementioned C18 column using methanol, acetonitrile, water, 0.05% PFPA and 0.01% FA and was operated at an overall higher organic content. Another aliquot was analysed using basic negative ion optimized conditions using a separate dedicated C18 column. The basic extracts were gradient eluted from the column using methanol and water, however with 6.5mM ammonium bicarbonate at pH 8. The fourth aliquot was analysed via negative ionization following elution from a HILIC column (Waters UPLC BEH Amide 2.1×150 mm, 1.7 µm) using a gradient consisting of water and acetonitrile with 10mM Ammonium Formate, pH 10.8. The MS analysis alternated between MS and data-dependent MSn scans using dynamic exclusion. The scan range varied slightly between methods but covered 70-1000 m/z.

### 2.5. Compound identification, quantification, and data curation

Raw data were extracted, peak-identified and QC processed using Metabolon’s hardware and software^[9]^. Compounds were identified by comparison to library entries of purified standards or recurrent unknown entities. Metabolon maintains a library based on authenticated standards that contains the retention time/index (RI), mass to charge ratio (m/z), and chromatographic data (including MS/MS spectral data) on all molecules present in the library. Furthermore, biochemical identifications are based on three criteria: retention index within a narrow RI window of the proposed identification, accurate mass match to the library ± 10 ppm, and the MS/MS forward and reverse scores between the experimental data and authentic standards. The MS/MS scores are based on a comparison of the ions present in the experimental spectrum to the ions present in the library spectrum.

Peaks were quantified using area-under-the-curve method. The raw area counts for each metabolite in each sample were normalized to correct for variation resulting from instrument inter-day tuning differences by the median value for each run-day, therefore, setting the medians to 1.0 for each run. This preserved variation between samples but allowed metabolites of widely different raw peak areas to be compared on a similar graphical scale.

### 2.6. Primary data

Metabolomic data project ID: CALI-020-0001. The non-targeted metabolomics analysis of 831 metabolites in 114 eligible subjects by UPLC-MS/MS method. Each biochemical in OrigScale is rescaled to set the median equal to 1. Then, missing values were imputed with the minimum. Exogenous substances (EDTA, molecular constructs, etc.) was removed. To minimise the effect of extreme numbers on the result, values outside mean ± 5SD were excluded.^[27]^.

### 2.7. Multivariate data analysis and pathway enrichment analysis

Multivariate data analysis was conducted using SIMCA (v.14.1; Umetrics, Umeå, Sweden). Orthogonal partial least squares-discriminant analysis (OPLS-DA) was used to increase the class separation, flatten dataset and locate variable importance in projection (VIP)^[30]^. The quality of models was validated using two parameters: R2Ycum (goodness of fit) and cumulated Q2 (Q2cum, goodness of prediction). A threshold of 0.5 is widely accepted in model classification to classify good (Q2cum ≥ 0.5) or poor (Q2cum < 0.5) predictive capabilities.

We validated the OPLS-DA model using a permutation test (200 times) to reduce the risk of overfitting and possibilities of false-positive findings. Plots show correlation coefficients between the original Y and the permuted Y versus the cumR2Y and Q2. Fitted regression lines were also displayed, which connects the observed Q2 to the centroid of the permuted Q2 cluster. The model was considered valid^[32]^ if (1) all Q2 values from the permuted data set to the left are lower than the Q2 value on the actual data set to the right and (2) the regression line has a negative value of intercept on the y-axis. To spot moderate and strong outliers, DModX test and Hoteling’s T-squared test were performed. These tests were performed using SIMCA. VIP values were then singled out by supervised investigations. VIP is a readout of the contribution of each variable on the X-axis to the model. It is summed over all components and weighted to the Y accounted for by every single component^[3]^. Therefore, VIP ranking reflects the metabolites’ contribution to the model. VIP > 0.5 indicate a higher percent variation explained when a metabolite is included in the model; VIP < 0.5 represent metabolite plays a less important role in it^[18]^. In this study, we select metabolites with VIP > 0.5 as major discriminant metabolites to expand our selection and pathway enrichment.

Pathway analysis and statistical analysis were performed using MetaboAnalyst 4.0^[7]^ with available Human Metabolome Database (HMDB) identifiers. For pathway analysis, metabolites were mapped to the Kyoto Encyclopedia of Genes and Genomes (KEGG)^[28]^ metabolic pathways, and quantitative pathway enrichment and pathway topology analysis were performed. To select the most discriminant pathways, we chose levels of impact and extremely significant difference (impact>0.3, *P*<0.05, respectively).

Lowess smoothing applied to plot an association between metabolites and the severity of ROP. And the resulting raw values were transformed to z scores using the mean and standard deviation.

### 2.8. Machine learning

Our simulation experiment was implemented in Python 3.7.7. We first use Chi-Squared (CHI2) to select important features. Random forest (RF) was further performed for classification. RF is a supervised classification technique based on an ensemble of decision trees^[5]^. We use the five-fold cross-validation to determine the optimal tuning parameters. The result was the average accuracy of the five-fold cross-validation procedure.

### 2.9. Statistical analysis

Demographic characteristics were first determined by QQ- and PP-plots. Normally distributed variables were presented as mean ± SD; otherwise, data were presented as median and mean ranks. Two-tailed paired Student’s t-Test and paired Wilcoxon tests were then used to compare variables whose distribution followed and did not follow a normal distribution, respectively, between ROP and non-ROP infants. Following log transformation and imputation of missing values, if any, with the minimum observed value for each compound, two-tailed paired Student’s t-Test was used to identify biochemicals that differed significantly between groups. Differences were considered significant when *P* < 0.05 as well as those approaching significance (0.05<*P*<0.10). The analysis was performed using SPSS, version 25.0. For all conditions, fold changes in metabolite levels relative to control group (non-ROP group) were calculated. Further statistical details can be found in the appropriate figure legends.

## Results

### 3.1. Demographic Characteristics

The demographic characteristics of the study cohort were presented in **Table 1**. Gestational age, birth weight and PMA of both study groups are strictly matched according to our matching criteria. Statistical differences are identified in delivery mode (cesarean section 33.30% in ROP vs 57.90% in control, *P* = 0.008), feeding strategy on the day (breast milk 43.90% in ROP vs 87.70% in non-ROP, *P* <0.001) and the amount of leukocyte in the blood ((8.61 ± 3.19) x10^9^/L in ROP vs (9.83 ± 3.08) x10^9^/L in non-ROP, *P* = 0.047) shows statistically significant differences. There is no statistical significance of differences in maternal age (*P* = 0.416), length of stay (*P* = 0.146), multiple pregnancy (*P* = 0.843) and sex (*P* = 0.570) between two groups. Interestingly, we did not find a statistically significant difference between two groups with regards to amounts of feeding, oxygen saturations or FiO2, oxygen inhalation mode on the day of the blood drawn. Previous history of administration of oxygen, switch of oxygen inhalation mode, sepsis and blood transfusion is comparative between ROP and control group. Similarly, no difference was found in concentration of C-reactive and the level of platelet between two groups. Noteworthy, none of them have received insulin infusion.

**Table 1.**
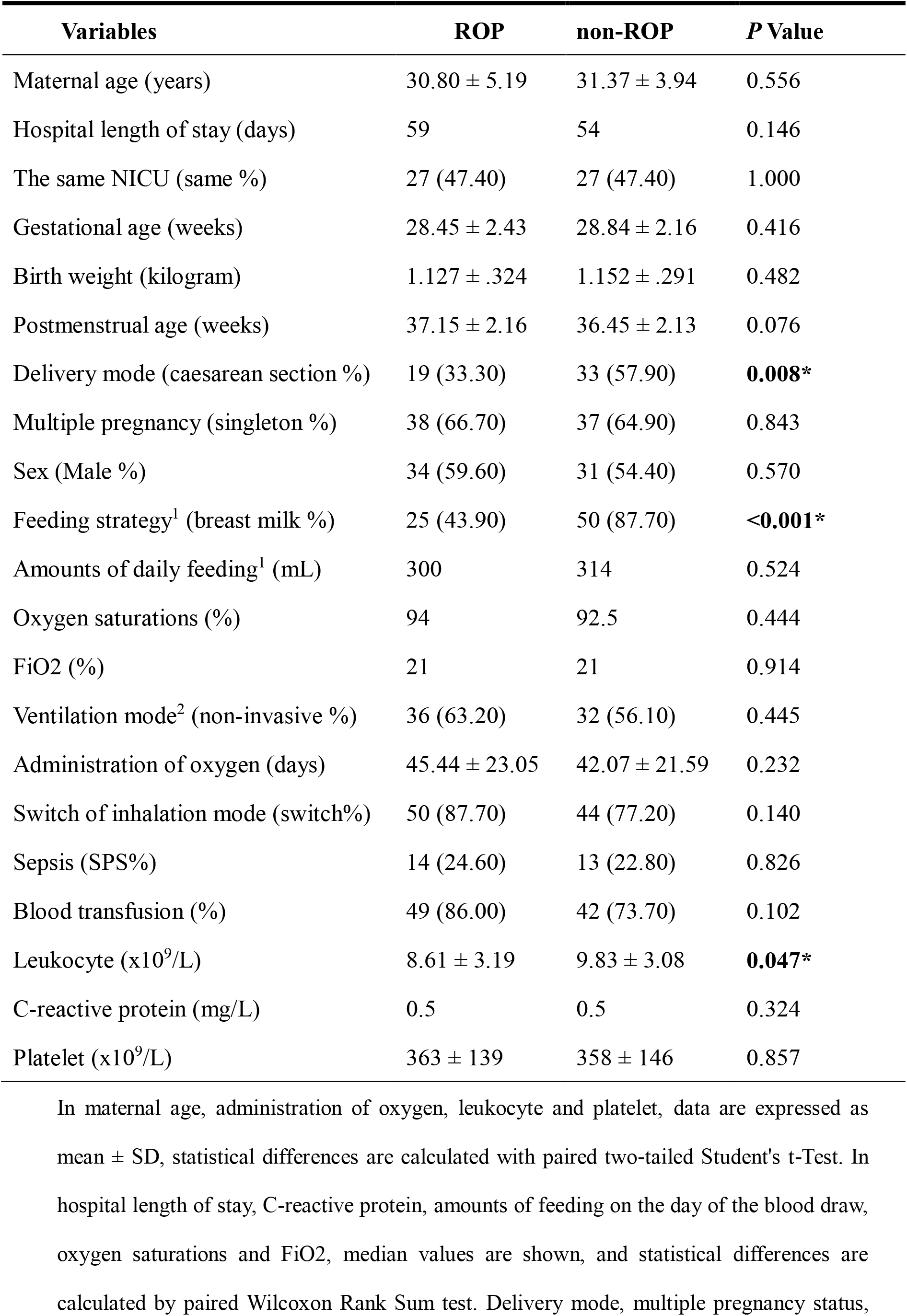
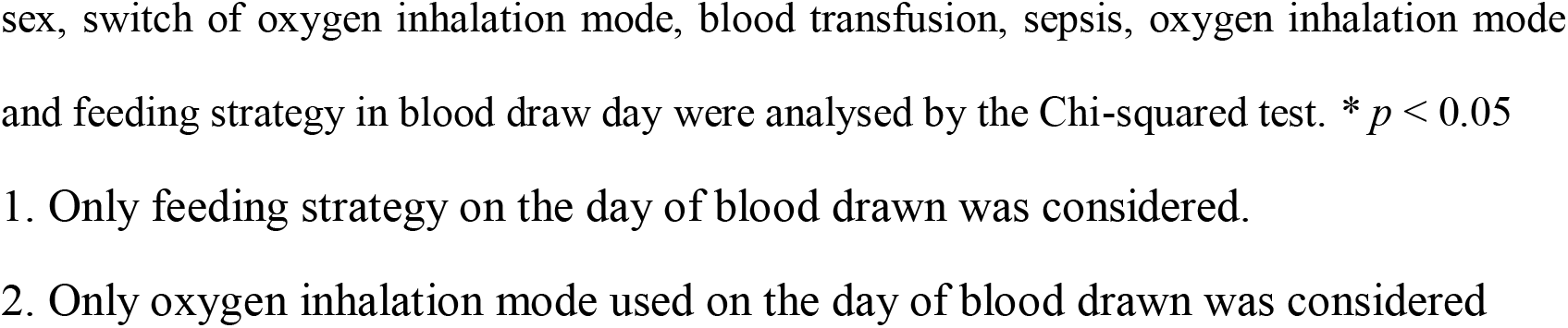
Demographic characteristics of participants.

| Variables | ROP | non-ROP | P Value |
| --- | --- | --- | --- |
| Maternal age (years) | 30.80 ± 5.19 | 31.37 ± 3.94 | 0.556 |
| Hospital length of stay (days) | 59 | 54 | 0.146 |
| The same NICU (same %) | 27 (47.40) | 27 (47.40) | 1.000 |
| Gestational age (weeks) | 28.45 ± 2.43 | 28.84 ± 2.16 | 0.416 |
| Birth weight (kilogram) | 1.127 ± .324 | 1.152 ± .291 | 0.482 |
| Postmenstrual age (weeks) | 37.15 ± 2.16 | 36.45 ± 2.13 | 0.076 |
| Delivery mode (caesarean section %) | 19 (33.30) | 33 (57.90) | **0.008\*** |
| Multiple pregnancy (singleton %) | 38 (66.70) | 37 (64.90) | 0.843 |
| Sex (Male %) | 34 (59.60) | 31 (54.40) | 0.570 |
| Feeding strategy<sup>1</sup> (breast milk %) | 25 (43.90) | 50 (87.70) | **<0.001\*** |
| Amounts of daily feeding<sup>1</sup> (mL) | 300 | 314 | 0.524 |
| Oxygen saturations (%) | 94 | 92.5 | 0.444 |
| FiO2 (%) | 21 | 21 | 0.914 |
| Ventilation mode<sup>2</sup> (non-invasive %) | 36 (63.20) | 32 (56.10) | 0.445 |
| Administration of oxygen (days) | 45.44 ± 23.05 | 42.07 ± 21.59 | 0.232 |
| Switch of inhalation mode (switch%) | 50 (87.70) | 44 (77.20) | 0.140 |
| Sepsis (SPS%) | 14 (24.60) | 13 (22.80) | 0.826 |
| Blood transfusion (%) | 49 (86.00) | 42 (73.70) | 0.102 |
| Leukocyte (x10<sup>9</sup>/L) | 8.61 ± 3.19 | 9.83 ± 3.08 | **0.047\*** |
| C-reactive protein (mg/L) | 0.5 | 0.5 | 0.324 |
| Platelet (x10<sup>9</sup>/L) | 363 ± 139 | 358 ± 146 | 0.857 |

In maternal age, administration of oxygen, leukocyte and platelet, data are expressed as mean ± SD, statistical differences are calculated with paired two-tailed Student's t-Test. In hospital length of stay, C-reactive protein, amounts of feeding on the day of the blood draw, oxygen saturations and FiO2, median values are shown, and statistical differences are calculated by paired Wilcoxon Rank Sum test. Delivery mode, multiple pregnancy status,
sex, switch of oxygen inhalation mode, blood transfusion, sepsis, oxygen inhalation mode and feeding strategy in blood draw day were analysed by the Chi-squared test. \* $p < 0.05$
1. Only feeding strategy on the day of blood drawn was considered.
2. Only oxygen inhalation mode used on the day of blood drawn was considered

### 3.2. Classification of metabolites

We firstly seek to categorize the total 742 metabolites pulled down from metabolomics analysis. **Figure 1** is a pie chart showing the fraction of eight main classes of metabolites, amongst which, most belongs to lipids (47.04%), followed by amino acids (29.11%). Whereas a relatively smaller fractions of metabolites are related to nucleotides (5.12%), cofactors and vitamins (4.99%), peptides (4.58%) and carbohydrates (4.04%). To compare the changing trend of these metabolite classes between control and ROP group, we constructed a heatmap (**Figure 2**). Most metabolites in lipids, nucleotides and carbohydrates categories are elevated in the ROP group, whereas those in amino acids, energy, cofactors and vitamins are fluctuated. It is worth mentioning that most xenobiotics occupy a small proportion of biochemicals that may indicate less external disturbance. **Supplementary Table 2** provides a summary of quantitative differences between the groups. There are 189 metabolites significantly differs, with 89 being elevated in the ROP and 100 lowered in the control. In addition, clear difference was also detected in another 67 metabolites, yet these do not achieve a level of statistical significance.

**Figure 1.**
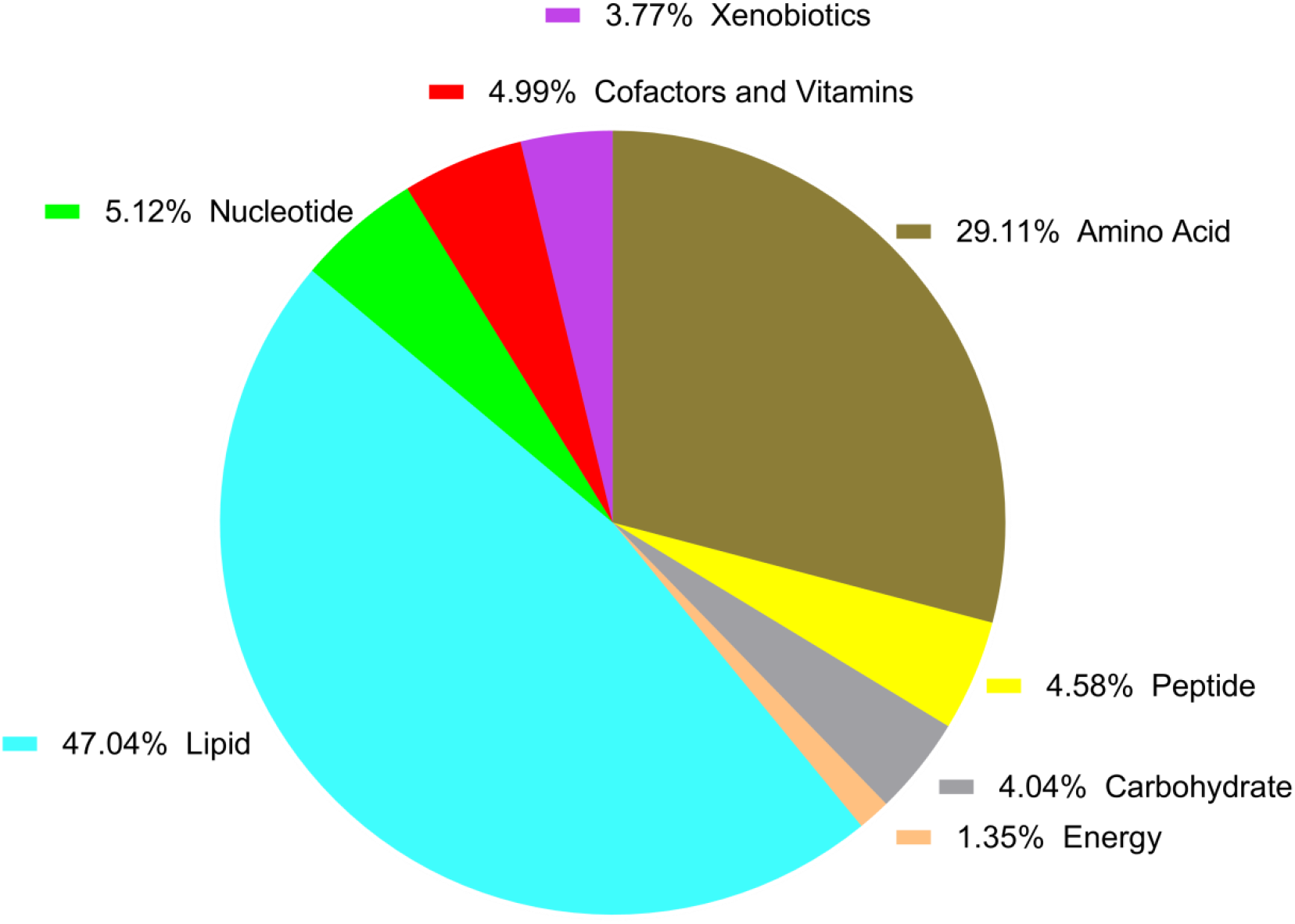
Pie chart showing fractions of main metabolite classes. Categories of metabolites are ordered from the highest to lowest fraction: lipids (47.04%), amino acids (29.11%), nucleotides (5.12%), cofactors and vitamins (4.99%), peptides (4.58%), carbohydrates (4.04%), xenobiotics (3.77%) and energy (1.35%).

**Figure 2.**
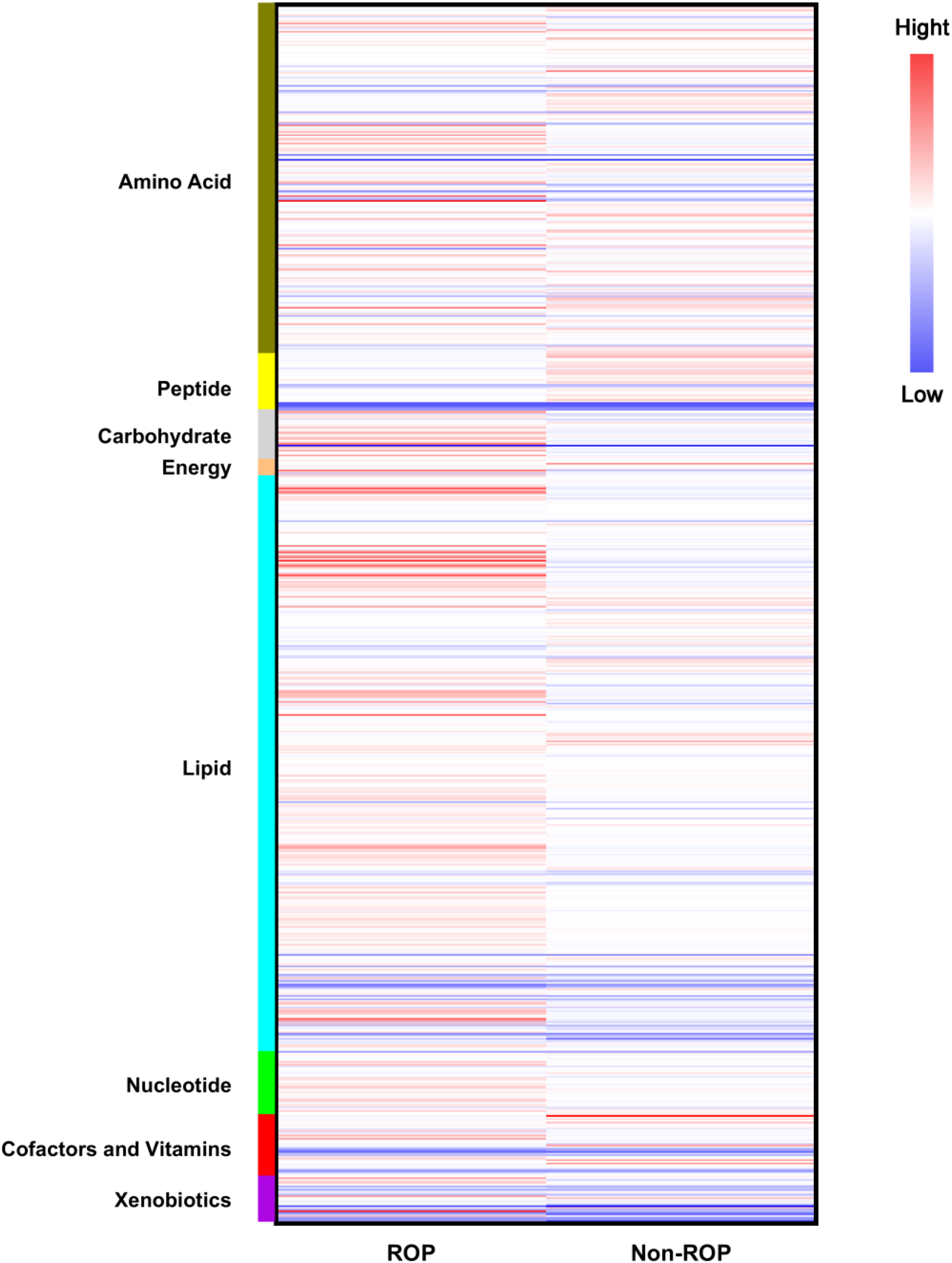
Heatmap showing metabolite categories in non-ROP and ROP group. The colour scales ranging from bright blue (low ratio) to bright red (high ratio) represent the relative ratio of Y=Log(Y)(intensity) between groups. In general, the level of metabolites in ROP slightly higher than that of non-ROP, especially in lipid.

### 3.3. Metabolic pathway enrichment analysis

#### 3.3.1. OPLS-DA model building and validation

We then investigate the OPLS-DA score plot (**Figure 3**) which revealed a clear and separate clustering between premature infants with and without ROP. Moreover, this OPLS-DA model achieves a R2Ycum of 0.908 and Q2cum of 0.523, meaning an excellent goodness of fit and a reliable predictive capacity.

**Figure 3.**
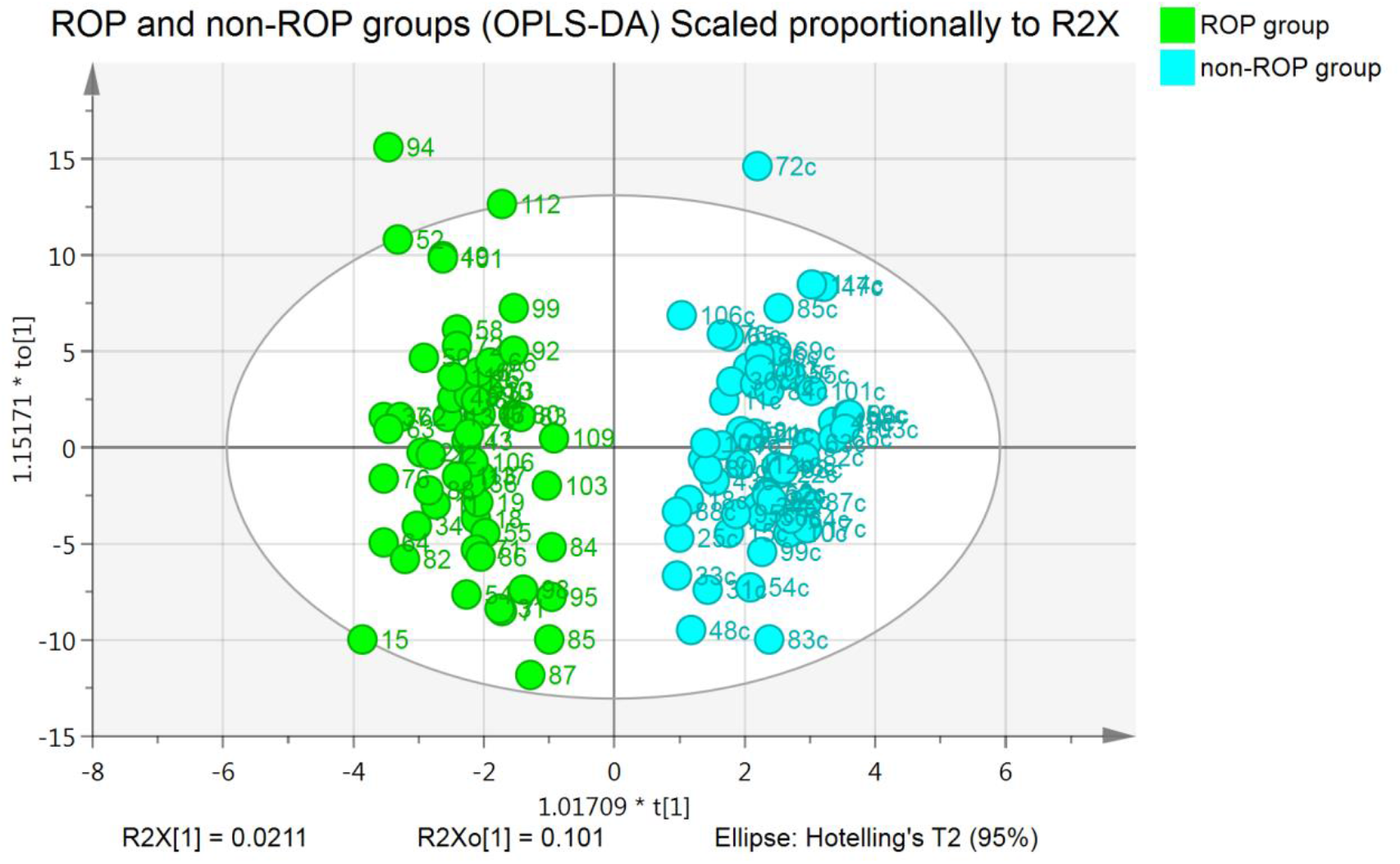
OPLS-DA scatter plot of samples from ROP and non-ROP groups. Samples from ROP and non-ROP group forms clearly separated clusters in the OPLS-DA analysis. R2Ycum = 0.908, Q2cum = 0.523. Green dots represent ROP (n = 57); blue dots represent non-ROP (n = 57).

By perform statistical validation of the corresponding OPLS-DA model by permutation testing (200 iterations) (**Supplementary Figure 1A**), we obtained all permuted R2s below or around 0.9 and most permuted Q2 below 0. Furthermore, all R2 and Q2 are lower than the original values on the right. Notably, Q2 regression line has a negative intercept at (0, -0.542). These collectively suggest that a valid model fitting which is unlikely to be built by chance. To identify strong and moderate outliers, we performed Hoteling’s T-squared test (**Supplementary Figure 1B**) and DModX test (**Supplementary Figure 1C**), respectively. No strong outliers in the sample can be identified, whereas infant 45 (in the ROP group) seems to show evident deviation in the DModX test.

#### 3.3.2. Contribution analysis of all metabolites in ROP

Based on the OPLS-DA model, we then construct an contribution plot, which ranks metabolites by their contribution to the model (**Supplementary Figure 2**). For clarity, only the top 78 metabolites are shown. The top three contributors are isocitric lactone, arabinose and mannose. To expand our pathway selection, there are 649 metabolites (VIP > 0.5) that were considered candidate to pathway enrichment.

#### 3.3.3. Pathway analysis

We use 649 metabolites with VIP values > 0.5 to carry out metabolite set enrichment analysis (MSEA) (**Figure 4**). Pathways at the top right achieve a high impact value and small *P* value (green box). We primary enriched yielded 9 pathways with obviously statistical significant difference (impact > 0.3, *P* < 0.05) (**Supplementary Table 3**), including arginine biosynthesis (-Log (*P*) = 11.584, Impact = 0.538), histidine metabolism (-Log (*P*) = 9.828, Impact = 0.533), glycine, serine and threonine metabolism (-Log (*P*) = 7.671, Impact = 0.729), phenylalanine, tyrosine and tryptophan biosynthesis (-Log (*P*) = 7.099, Impact = 1.000), alanine, aspartate and glutamate metabolism (-Log (*P*) = 6.832, Impact = 0.741), phenylalanine metabolism (-Log(p) = 5.936, Impact = 0.619), beta-alanine metabolism (-Log (*P*) = 5.373, Impact = 0.672), taurine and hypotaurine metabolism (-Log (*P*) = 5.306, Impact = 1.000), arginine and proline metabolism (-Log (*P*) = 3.955, Impact = 0.376).

**Figure 4.**
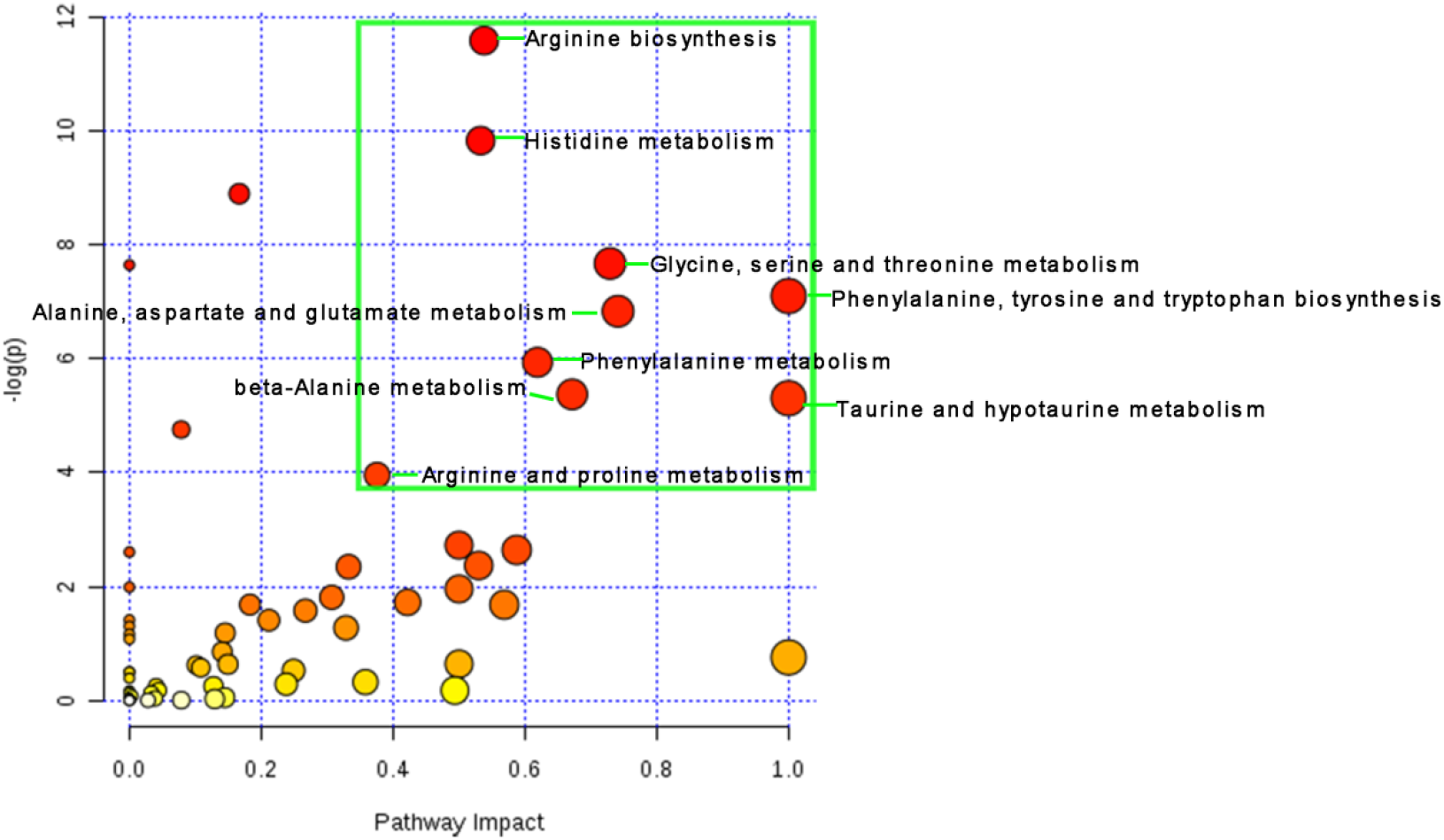
Pathway enrichment results. Pathway impact is determined by topological analysis (x-axis) and enrichment log (*P*) value is adjusted by the original *P* value (y-axis). Node colour is based on its *P*-value, and the node size is based on pathway impact values. The top nine most significantly affected metabolic pathways (Impact > 0.3, *P* < 0.05) are inside the green box.

### 3.4. Feature selection and random forest analysis

To narrow down and identify the most optimal biomarker combination that are useful in differentiating ROP from non-ROP infants, we implemented 742 metabolites and then use the chi-square feature selection based random forest method (**Figure 5**). We obtain an unbiased estimate of the predictive performance of the random forest model which typically outputs the top 30 metabolites, the top six are isocitric lactone (variable importance 0.0567); creatinine (0.0416); methylsuccinate (0.0413); arabinose (0.0405); arginine (0.0359) and ribitol (0.0348). However, only differences in creatinine; ribitol; orotidine; cis-4-decenoate (10:1n6); 3-(4-hydroxyphenyl) lactate; undecanoate (11:0); kynurenine and glutamate, gamma-methyl ester achieve statistical significance (*P* < 0.05). Ornithine; 10-undecenoate (11:1n1) and picolinoylglycine have the difference approaches statistical significance (0.05 < *P* < 0.10).

**Figure 5.**
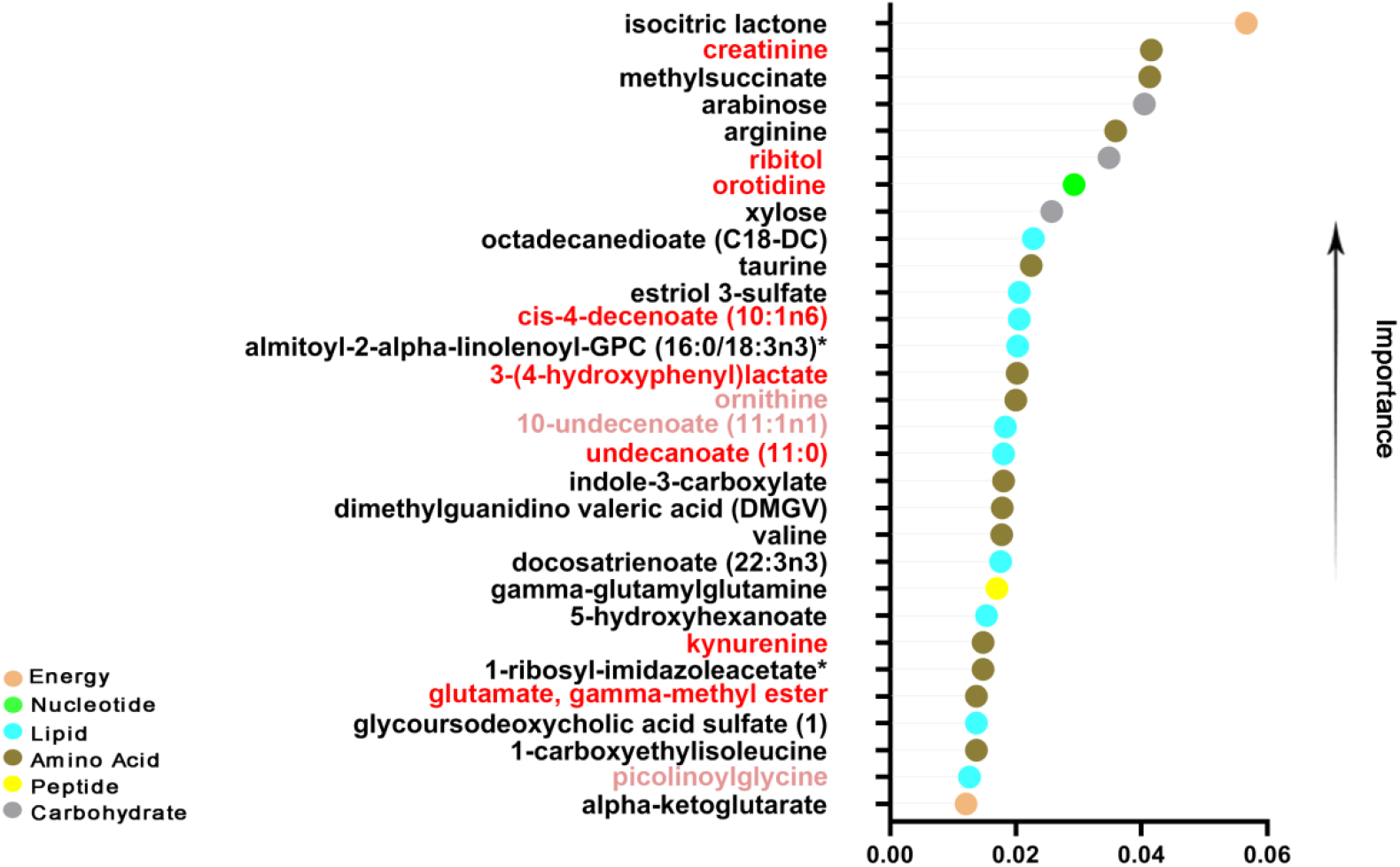
Random forest graph. The vertical axis shows the top 30 metabolites and the horizontal axis shows the corresponding importance of the feature. Different colours represent different metabolites classes. Isocitric lactone has the highest feature importance.

From the random forest and the pathway enrichment results, we identify 6 pathways corresponding to 9 metabolites as the top 30 most significantly affected metabolites in the ROP group (**Figure 6**). In the alanine, aspartate and glutamate metabolism, the average concentration of glutamate, gamma-methyl ester are increased in ROP group compared to controls. This indicates a shift in the redox homeostasis and fatty acid oxidation (FAO), suggesting a more antioxidative state. However, in arginine biosynthesis and arginine and proline metabolism, the average level of ornithine and DMGV is reduced in ROP, whereas arginine is slightly elevated. In addition, in phenylalanine, tyrosine and tryptophan biosynthesis, the indole-3-carboxylate is increases and 3-(4-hydroxyphenyl) lactate and kynurenine shows a decreasing trend in the ROP group.

**Figure 6.**
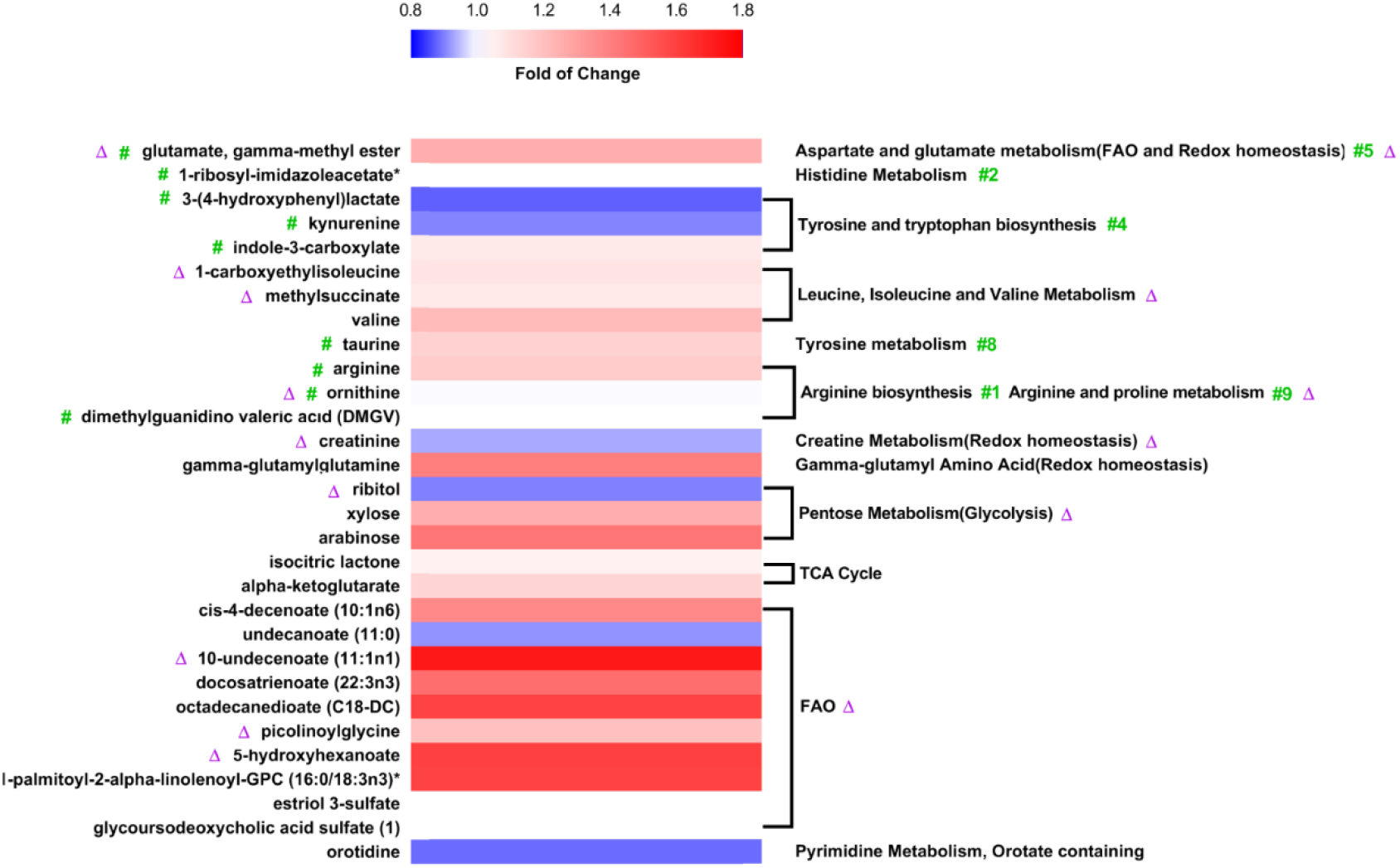
Correlation heatmap of the top 30 metabolites and pathways selected from random forest analysis. The green wells on the left denote metabolites that corresponding to enriched metabolic pathways (on the right with green colour #1, #2, #4, #5, #8 and #9). Statistically significant differences are indicated by a purple triangle on the left and right (adjusted *P* < 0.05). Interestingly, arginine biosynthesis, FAO and redox homeostasis are the results of enriched pathways and both before and after adjustment.

After adjusting for statistically significant differences (delivery mode, feeding strategy and leukocyte), the following metabolites of the first 30 metabolites in random forest remain statistically significant. Creatinine, ribitol and glutamate, gamma-methyl ester that are still independent risk factors for ROP (**Table 2**). The higher the value of creatinine, the lower the risk of ROP, indicating it is a strong protective factor (P = 0.038, Wald = 4.308, odds ratio [OR] = 8.1465E-7, confidence interval [CI] = (1.4484E-12, 0.458)). Ribitol is a weak protective factor for ROP (*P* = 0.016, Wald = 5.770, OR = 0.018, CI = (0.001, 0.476)). The higher the level of glutamate, gamma-methyl ester the higher the risk of ROP, suggesting it being a strong risk factor (*P* = 0.015, Wald = 5.908, OR = 1.6491E+05, CI = (10.238, 2.6563E + 09)). Notable metabolites which close to statistical significance (0.05 < *P* < 0.1) show a strong statistically significant after adjusting, namely, ornithine, 10-undecenoate (11:1n1) and picolinoylglycine. Therefore, we reasoned that creatinine; ribitol; glutamate, gamma-methyl ester; ornithine; 10-undecenoate (11:1n1) and picolinoylglycine are potential biomarkers of ROP (**Figure 7**). Taken together the results from **Figure 6**, it appears that pathways related to glycometabolism, redox homeostasis, lipid metabolism and arginine, tyrosine and tryptophan metabolic pathway are likely to be closely associated with ROP.

**Figure 7.**
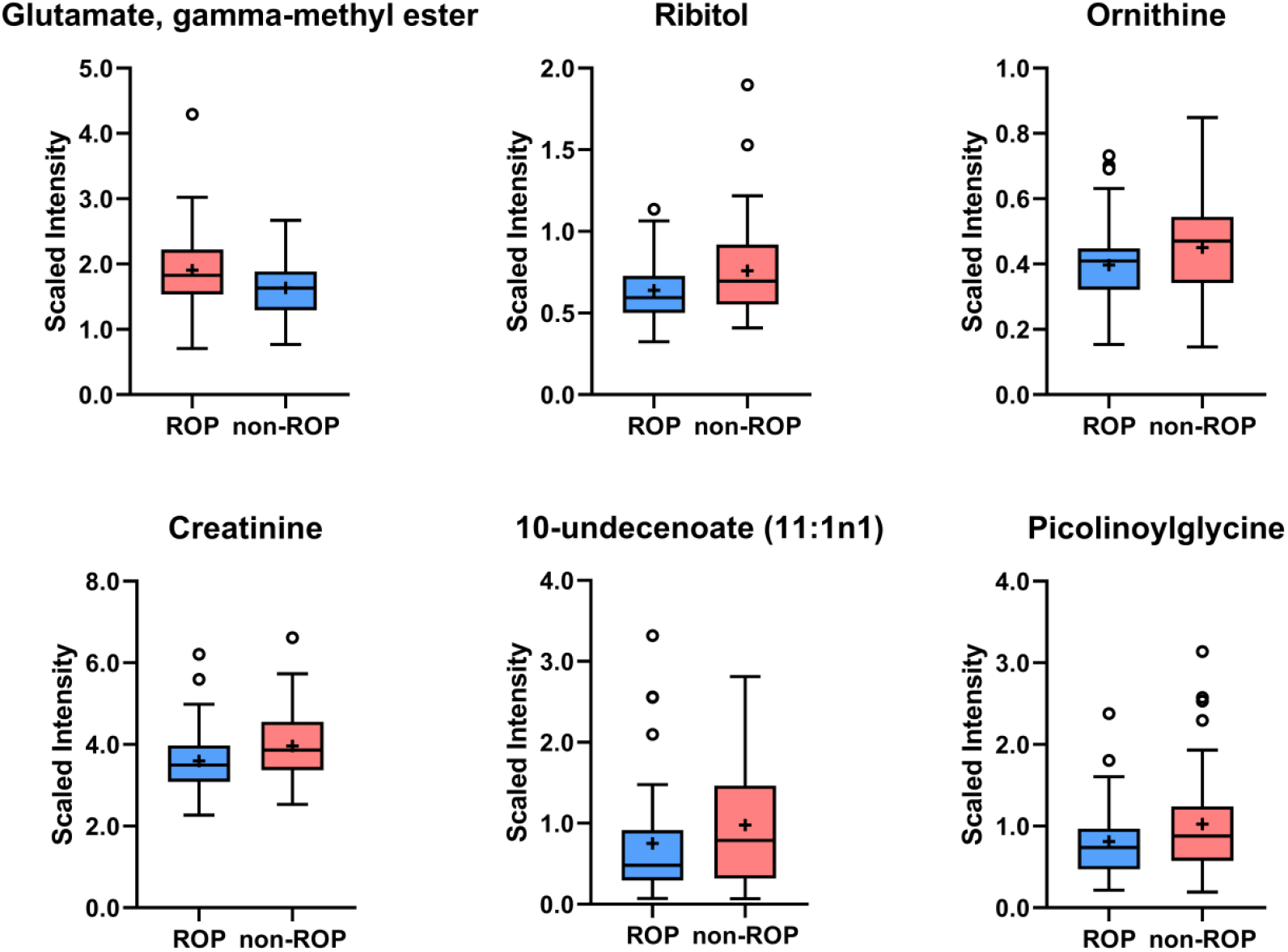
Box plot of potential biomarkers of ROP. Creatinine, ribitol and glutamate, gamma-methyl ester shows statistical difference (*P* < 0.05); ornithine, 10-undecenoate (11:1n1) and picolinoylglycine approach statistical significance (0.05 < *P* < 0.1). Light red represents elevated levels of metabolite, and light blue means reduced levels of the metabolites. Circles represent outliers and the plus signs serve as the mean.

**Table 2.**
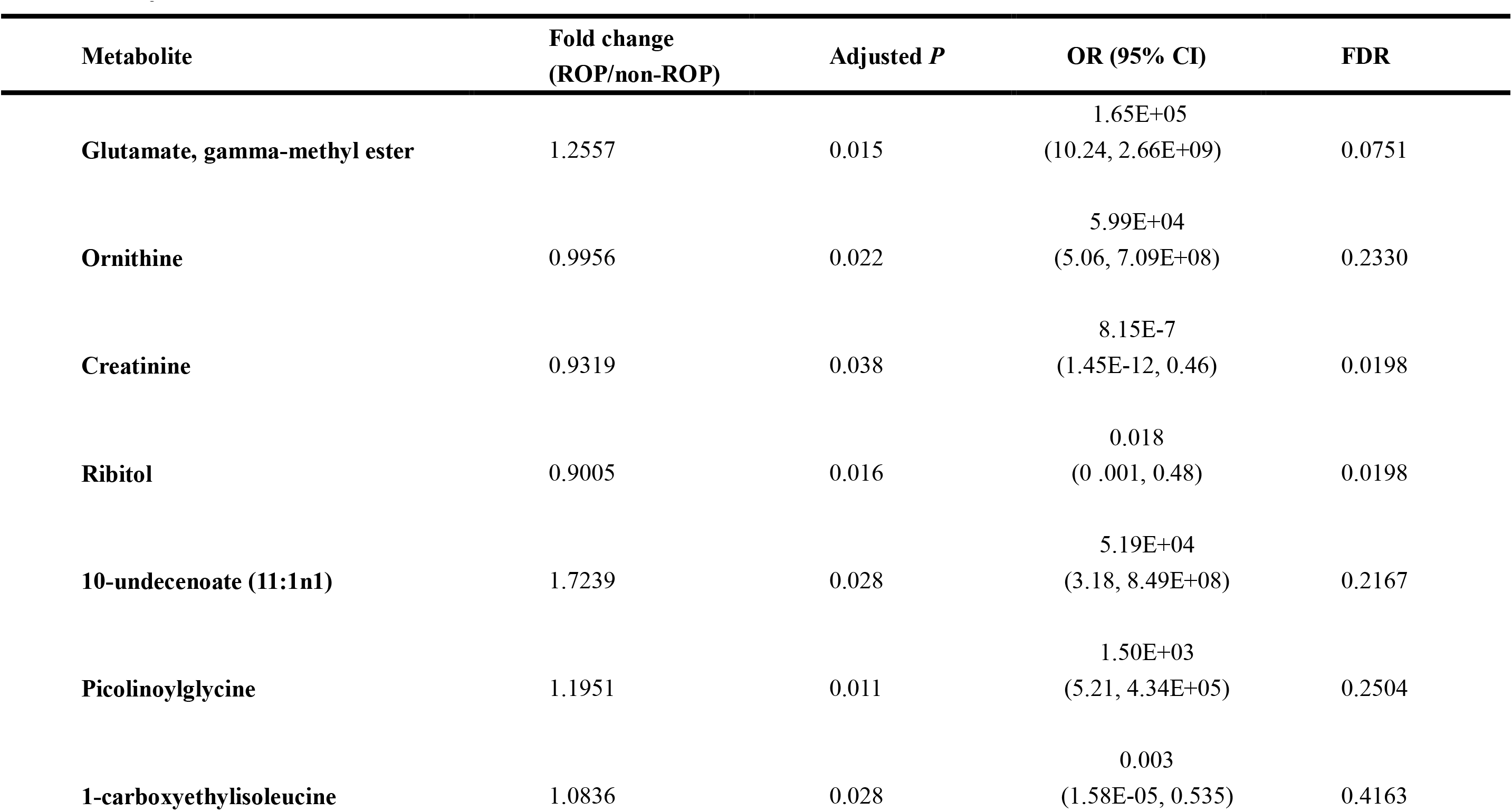
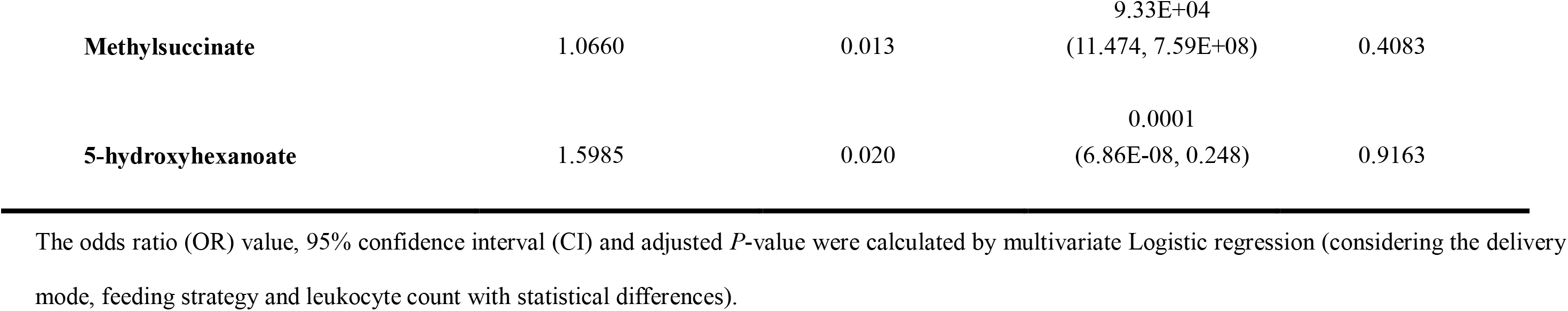
Adjusted metabolites with statistical differences.

| Metabolite | Fold change<br>(ROP/non-ROP) | Adjusted *P* | OR (95% CI) | FDR |
| --- | --- | --- | --- | --- |
| Glutamate, gamma-methyl ester | 1.2557 | 0.015 | 1.65E+05<br>(10.24, 2.66E+09) | 0.0751 |
| Ornithine | 0.9956 | 0.022 | 5.99E+04<br>(5.06, 7.09E+08) | 0.2330 |
| Creatinine | 0.9319 | 0.038 | 8.15E-7<br>(1.45E-12, 0.46) | 0.0198 |
| Ribitol | 0.9005 | 0.016 | 0.018<br>(0 .001, 0.48) | 0.0198 |
| 10-undecenoate (11:1n1) | 1.7239 | 0.028 | 5.19E+04<br>(3.18, 8.49E+08) | 0.2167 |
| Picolinoylglycine | 1.1951 | 0.011 | 1.50E+03<br>(5.21, 4.34E+05) | 0.2504 |
| 1-carboxyethylisoleucine | 1.0836 | 0.028 | 0.003<br>(1.58E-05, 0.535) | 0.4163 |
| **Methylsuccinate** | 1.0660 | 0.013 | 9.33E+04<br>(11.474, 7.59E+08) | 0.4083 |
| **5-hydroxyhexanoate** | 1.5985 | 0.020 | 0.0001<br>(6.86E-08, 0.248) | 0.9163 |

The odds ratio (OR) value, 95% confidence interval (CI) and adjusted *P*-value were calculated by multivariate Logistic regression (considering the delivery mode, feeding strategy and leukocyte count with statistical differences).

### 3.5. Biomarkers for ROP

We then investigate the ROC plot to assess how good are the above chosen metabolic biomarkers in predicting ROP (**Supplementary Figure 3**). AUC of potential biomarkers: glutamate, gamma-methyl ester (AUC = 0.642, CI = (0.541, 0.743)), ornithine (AUC = 0.671, CI = (0.5719, 0.7694)), creatinine (AUC = 0.572, CI = (0.467, 0.677)), ribitol (AUC = 0.766, CI = (0.6783, 0.8536)), 10-undecenoate (11:1n1) (AUC = 0.602, CI = (0.497, 0.707)), picolinoylglycine (AUC = 0.618, CI = (0.514, 0.722)) are greater than 0.50. Notably, the AUC of ribitol is even greater than 0.766, indicating its reliable predictive ability. According to the hypothesis test (H0 = 0.5), and after adjusting for confounding factors creatinine, ribitol, glutamate, gamma-methyl ester, ornithine, 10-undecenoate (11:1n1) and picolinoylglycine are confirmed to be true positive and do not seem to be obtained randomly (**Table 2**). In particular, the accuracy of ribitol - based diagnosis is the best (AUC = 0.766, sensitivity = 71.9%, and specificity = 71.9%) amongst all potential discriminant metabolites.

To investigate correlations between level of potential biomarkers with severity of ROP, we employ the proportional odds model with ordinal logistic regression. The result of parallel lines test suggests a valid proportional odds assumption (χ^2^ = 12.892, *P =* 1.000). In goodness-of-fit tests, the deviance test indicated the model fits well (χ^2^ = 229.829,*P =* 1.000), which is better than the model with only a constant (χ^2^ =40.073,*P* < 0.001). we observe that the level of creatinine and ribitol decreases with increased ROP severity (*P =* 0.018, OR = 0.027, 95%CI = (0.001,0.544); *P =* 0.011, OR = 0.448, 95%CI= (0.241,0.834), respectively). However, we do not identify a clear association between glutamate, gamma-methyl ester; ornithine; 10-undecenoate (11:1n1) or picolinoylglycine with ROP severity after adjusting for delivery mode, feeding strategy and leukocyte. Our study provides the very first data indicating a potential association between declined creatinine/ribitol and the severity of ROP. Nonetheless, the Lowess curves of creatinine (first rises and then falls) does not perfectly predict the ROP severity as it did in the proportional odds model, this may be attributed to coarse grading systems, extreme outliers or small number of severe ROP infant (2 infants of AP ROP and 1 infants of stage 4 ROP) (**Supplementary Figure 4**).

Furthermore, we also find that premature with more severe ROP is related to be fed with a combination of breast milk and parenteral nutrition (5.9-fold, 95% CI = ( 2.564, 13.699), χ^2^ = 17.315, *P <* 0 .001), higher spontaneous labour rate (2.3-fold, 95%CI = (1.034, 5.115), χ^2^ = 4.168, *P =* 0 .041) and higher level of leukocytes (with every 1 (x10^9^/L) increase, the risk of ROP is elevated by 1.1%, 95% CI = (1.001, 1.295),χ^2^ = 3.928, *P =* 0 .048). These findings are consistent with our previous study on targeted analysis of blood spot samples from ROP infants^[53]^. The reason for a higher fraction of ROP infants born via spontaneous labour may be due to those underwent prolonged labour are more likely to be exposed to lengthened anoxia, which exaggerate fluctuation of oxygen during delivery.

Beyond this, none of the above 6 potential biomarkers (glutamate, gamma-methyl ester, ornithine, creatinine, ribitol, 10-undecenoate (11:1n1) and picolinoylglycine) were linearly correlated with the change of PMA, BW, and GA by using simple linear regression analysis. (ROP group: R = 0.025, F = 1.43, *P* = 0.237; R = 0.003, F = 0.16, *P* = 0.689; R = 0.012; F = 0.68, *P* = 0.413; R = 0.019; F = 1.04, *P* = 0.312; R = 0.041; F = 2.38, *P* = 0.129; R = 0.011; F = 0.63, *P* = 0.430, respectively; non-ROP group: R = 0.005; F = 0.29, *P* = 0.592; R = 0.007; F = 0.38, *P* = 0.539; R = 0.016; F = 0.89, *P* = 0.350; R = 0.006; F = 0.32, *P* = 0.576; R < 0.001; F = 0.01, *P* = 0.920; R = 0.025; F = 1.38, *P* = 0.245, respectively). We plotted the second order polynomial fits to serve as a correlation between biomarkers and PMA (**Figure 8).** Our study indicating a potential association between glutamate, gamma-methyl ester, ornithine, creatinine, ribitol, 10-undecenoate (11:1n1), picolinoylglycine and risk/severity of ROP, but not the change of PMA, BW and GA.

**Figure 8.**
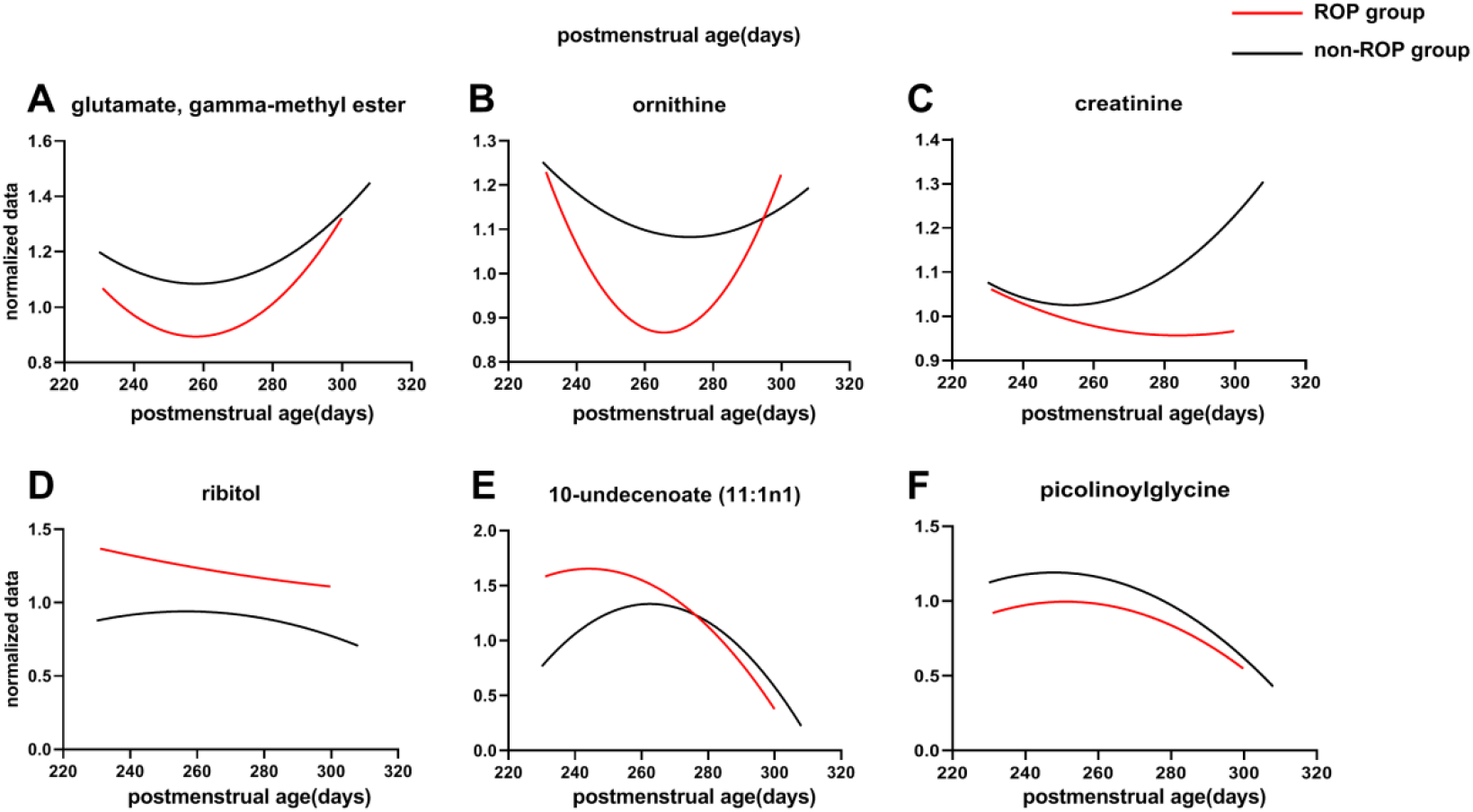
Non-linear regression analysis correlation between PMA(days) and biomarkers. The curve was obtained by fitting to a two-parameter model in non-linear regression analysis. Potential biomarkers (glutamate, gamma-methyl ester, ornithine, creatinine, ribitol, 10-undecenoate (11:1n1), and picolinoylglycine) would presumably vary with PMA (days), however, they did not verify the predicted linear relationship. Red lines represent ROP group; black lines represent non-ROP group.

## Discussion

In the present study, we compared the serum metabolomic profiling between strictly matched control and ROP premature infants. Analysis revealed a few altered metabolic pathways and identified 6 major potential biomarkers of ROP, which are reliable at predicting the development and severity of ROP. We will discuss these biomarkers and relevant pathways to illustrate the rationale of them being predictive of ROP. We constructed a comprehensive map of potential pathways identified in the present study to the development of ROP (**Figure 9**).

**Figure 9.**
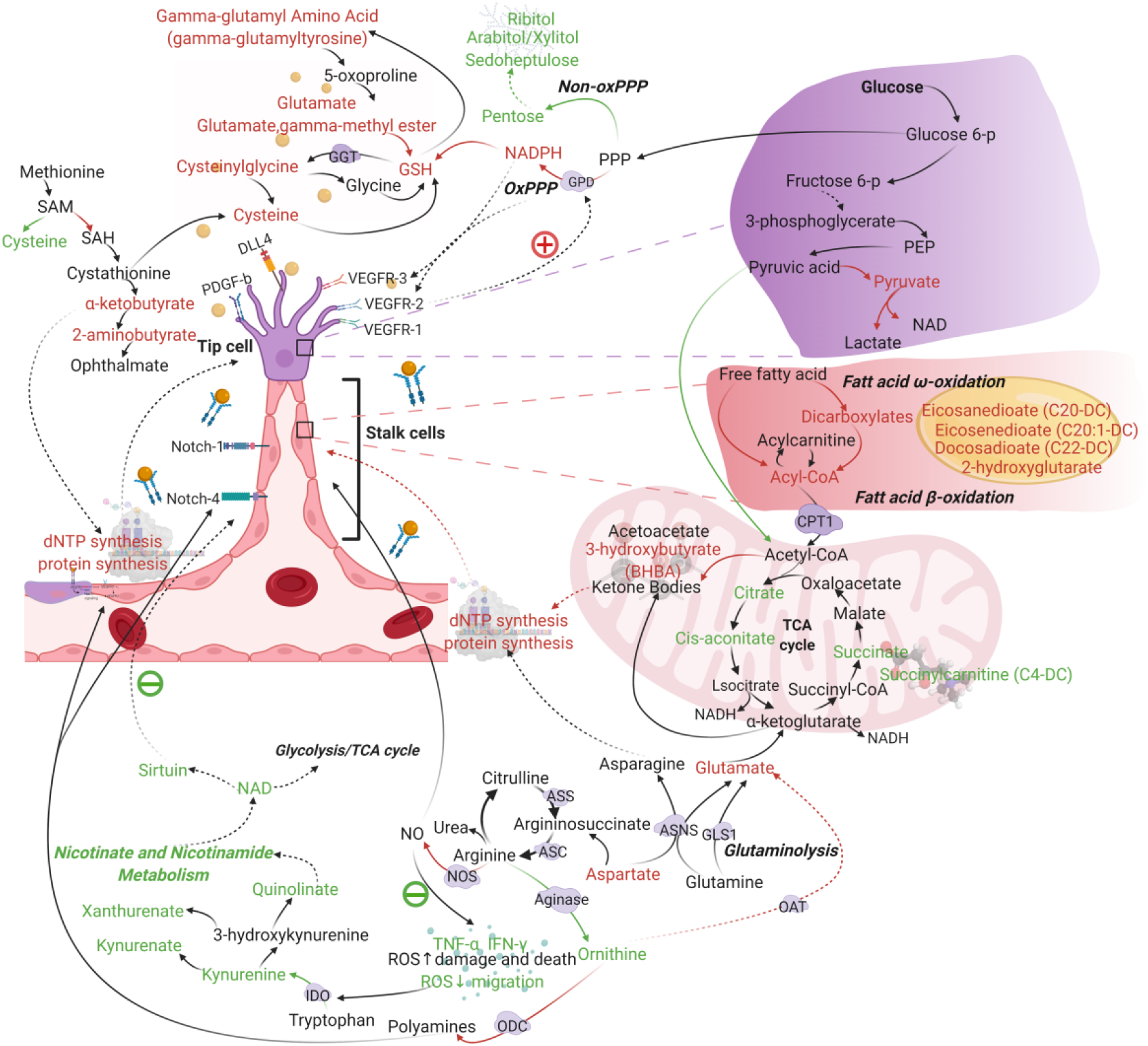
Schematics of potential pathways associated to ROP development. Red words, increased metabolite in ROP group; Green words, decreased metabolite in ROP group. Red arrows indicate pathways that are more active reaction and green arrows represent a less-active reaction. Purple background denotes key enzymes of relevant pathways. Abbreviations: CPT1 = carnitine palmitoyltransferase 1; PEP = phosphoenolpyruvic acid; ASNS = asparagine synthase; GLS1 = glutaminase 1; GPD = glucose-6-phosphate dehydrogenase; GSH = glutathione; GGT = gamma-glutamyl transferase; SAM = S-adenosylmethionine; SAH = S-adenosylhomocysteine; ASS = argininosuccinate synthase; ASL = argininosuccinate lyase; NOS = nitric oxide synthase; IDO = indoleamine-2,3-dioxygenases.

### 4.1. Glycometabolism and redox homeostasis

ROP is characterized by its vascular abnormality, such as neovascularization. Increased endothelial sprouting and proliferation are major cellular events causing pathological proliferative retinopathies, therefore, deciphering the molecular mechanisms underlying these early cellular events are key to understanding and further developing novel therapeutic approaches for the prevention or treatment of ROP. Our discussion will also focus on the implication of identified metabolite changes in the major vasculature component (i.e., endothelial cells).

The retina is one of the most energy-demanding tissues of the body. Under hypoxic conditions, there are still a considerable fraction (80-96%) of glucose being converted to lactate^[24, 25]^. Similar to the retina, ECs produce adenosine triphosphate (ATP) by aerobic glycolysis regardless of whether oxygen is present which referred to as the Warburg effect^[11]^. Aerobic glycolysis is rapidly increased to supplement cellular energy needs (within seconds to minutes), while oxidative phosphorylation (OXPHOS) is 100 times slower than that of aerobic glycolysis^[44]^. These make prompt metabolism to adapt to booming demand on the energy needs, as long as ECs have received stimulation of VEGF. Yizhak et al^[54]^. also confirmed that ECs migration utilizes aerobic glycolysis to generates ATP rather than OXPHOS.

In the current study, we found that the level of glycolytic intermediates, pyruvic acid, pyruvate and lactate, were higher in ROP infants’ serum, while the tricarboxylic acid cycle (TCA cycle) metabolites, such as citrate, aconitate and succinylcarnitine (C4-DC), is lower. Such differences are indicative of perturbations in aerobic oxidation. In other words, it demonstrates that aerobic glycolysis is over-active in infants with ROP. In fact, these observations were in accordance with the exuberant proliferation of neovascularization endothelial cells in ROP. In addition, a small amount of mitochondrial pyruvate carrier is responsible for the transport of pyruvate from glycolysis to mitochondria for OXPHOS, which decrease of would also lead to a series of retinal diseases^[20]^.

Increased level of phosphate pentose pathway (PPP) and hexosamine biosynthesis pathway (HBP) provide another line of evidence supporting that the aerobic glycolysis is more active in ROP. Glucose-6-phosphate enters the PPP metabolic pathway under the action of glucose-6-phosphate dehydrogenase (GPD) to promote the synthesis of ribose-5-phosphate, which is necessary for nucleotide biosynthesis^[35]^. PPP is composed of oxidized (oxidative branch, oxPPP) and non-oxidized branches (non-oxidative branch, non-oxPPP), the former plays an influential role in the activity and migration of EC^[2, 33]^. OxPPP is facilitated by GPD, whose is controlled by VEGF^[13, 34]^. In redox homeostasis, ROP group presented elevated levels of cysteine, alpha-ketobutyrate, 2-aminobutyrate, glutathione, glutamate and gamma-methyl ester, which can lead to decreased production of relevant metabolites, such as creatinine. These alternations can also relate to an increase in nicotinamide-adenine dinucleotide phosphate (NADPH) from the oxPPP branch, which maintands redox state of GSH^[34]^, such state is important in inhibiting the production of reactive oxygen species. It was shown in OIR mice studies that a considerable amount of reactive oxygen species produced by NADPH was clearly associated with retinal neovascularization^[12, 45, 47, 49]^. Consistent with increased GSH levels, we found an accumulation of cysteinylglycine in the ROP compared to control, suggesting alteration in gamma-glutamyl transferase (GGT) activity. However, no significant change was identified for gamma-glutamyl amino acids (except for gamma-glutamyltyrosine). In addition, we detected downregulation in several aminosugars, such as N-acetylneuraminate and erythronate. These can be converted from matabolites in the PPP pathway, for instance, ribitol, arabitol/xylitol and sedoheptulose, which were declined in the ROP group. This may also account for that oxPPP and non-oxPPP are interconvertible in the PPP pathway ^[2]^ (**Figure 9**).

In summary, these results suggest a disruption in glycometabolism and redox inbalance in the serum of ROP infants. This led to an accumulation of correlated metabolites, such as NADPH, GSH and hexosamine, together with a decrease TCA cycle-related molecule. Our prediction model also suggests that, creatinine, ribitol, glutamate and gamma-methyl ester are likely to be potential biomarkers for ROP.

### 4.2. Lipid metabolism pathway

Compared to control, we found that FAO process in the ROP group is highly active, this is revealed by increased level of most fatty acids, for instance malonylcarnitine (C3DC) which may lead to impaired angiogenesis^[6, 40]^. In addition, in the present study we found the level of aspartic acid (nucleotide precursor) and glutamate were also increased in the ROP group. These two amino acids are known to complement each other to produce α-ketoglutarate and maintain protein and/or nucleotide biosynthesis needed for vascular endothelial cell proliferation^[23]^. In accordance with the present results, our previous targeted metabolomics study reported a similar change in the C3DC and glutamate in the ROP group^[53]^..

Corresponding to this observation, in omega-oxidation, we also detected elevations in dicarboxylic fatty acids, eicosanedioate (C20-DC) and docosadioate (C22-DC, trending). Although beta-oxidation represents the primary route of fatty acid metabolism, if this process is overwhelmed or impaired, dicarboxylate-generating omega-oxidation (i.e. oxidation of the terminal carbon of fatty acid in peroxisomes and endoplasmic reticulum) may be utilized to help meet energy demand. Several lines of evidence suggest that suppressed carnitine palmitoyltransferase 1A can induce retinal vascular deficiency and reduce pathological angiogenesis in a model of ROP^[40]^.

ECs rely heavily on aerobic glycolysis^[12]^, whereas FAO provides only contributes to 5% of total ATP production in ECs^[8]^. Schoors et al.^[40]^ proposed that this would ensure ECs have enough *de novo* synthesis of deoxynucleotides during angiogenic sprouting. It also enables ECs to selectively regulate the proliferation, instead of migration, of ECs via FAO. This is consistent to the idea that endothelial migration mainly depends on glycolysis^[54]^, which suggested that different metabolic pathways regulate different ECs functions in the process of vascular sprouting. Furthermore, Egnatchik et al. demonstrate that systemic FAO blockade with a chemical inhibitor alleviates excessive angiogenesis in a mouse model of retinopathy of prematurity^[38]^. With regards to the FAO process, our analysis suggests that 10-undecenoate (11:1n1) and picolinoylglycine are potential biomarkers for ROP.

### 4.3. Arginine metabolism pathway

In this pathway, we found a significantly lower level of ornithine in the ROP group. This is likely to be related to reduced arginase activity and/or the higher ornithine decarboxylase and/or ornithine-oxo-acid transaminase activity. It has been shown that the up-regulation of arginase activity is associated with inflammation, oxidative stress and peripheral vascular dysfunction^[41]^.

These finding is in agreement with the early untargeted metabolomics studies performed in OIR by Lu et al^[15]^ and plasma metabolites research of ROP infant by Zhou et al^[55]^. However, their baseline data are not comprehensive, especially lack the consideration of feeding strategy and administration of oxygen on plasma metabolites, thus the results are not reliable. Besides, results were inconsistent without adjustment confounding factors, and, the method they used is defective (not untargeted, globally accepted standard regimen). What is more, the sample size is too small which will lead to instability. Without mechanism inference, it is neither a network nor a comprehensive view to analyze the possible role of metabolic pathways in the ROP. In the present study, we used a rigorous matching strategy using detailed demographic and clinical information are shown in **Table 1.**, whereas others used self-reported data or less stringent matching. To make our results more reliable and scientific, we use Metabolon, Inc. platform and multiple approaches such as machine learning and multivariate data analysis after adjustment. Therefore, we gave reasonable speculation to several potential pathways for the results.

Furthermore, arginase has been recognized as a therapeutic target for CNS disorders and cardiovascular disorders for its role in mediating damage in the retinal neovascularization and targeting early phase of neovessle growth^[36, 39]^. In mammalian cells, nitric oxide (NO) is known to regulate angiogenesis, neurotransmissions, immune responses, oxidative stress, and activate endothelial progenitor cells^[16, 39]^. Arginine is a substrate of NO synthase (NOS) and arginases, these two enzymes counterbalance each other. Therefore, NOS will be activated when arginase activity is inhibited. In addition, during OIR ischemia phase, when arginase activity is downregulated, an increase in iNOS expression, physiologic angiogenesis, vitreoretinal neovascularization and impaired retinal function were reported^[42]^. Furthermore, Wang et al.^[48]^ also found that decreased ROS level led to angiogenesis and ECs migration, which is likely to be associated with neovascularization in ROP.

On the other hand, increased ornithine decarboxylase activity could lead to excessive production of proline and polyamines, causing collagen deposition, pathological neovascularization and vascular fibrosis^[51]^. Moreover, proline is indispensable for the synthesis and maturation of collagen, which is necessary for the maintenance of blood vessel integrity and angiogenesis; additionally, polyamines are vital factors for cell proliferation, ion channel function and neuroprotection^[29]^. Therefore, upregulated ornithine decarboxylase activity may be a potential contributor for fibrovascular proliferation and retinal detachment at the late stage of ROP.

### 4.4. Limitations of the study

This study is to some extent limited. First, in order to adhere to our strict matching criteria while facing the number of cases diagnosed in south China, the case number is relatively small. while wring this manuscript, we are planning a nationwide and multi-ethnic study, in the hope of acquiring more cases and obtaining a wider scope. Second, to understand the causal roles of metabolic biomarker and disease development and/or progression, one should ideally develop prospective longitudinal studies that allow future disease risks to be evaluated on the basis of multi-omics information or other tissue, such as urine and faeces. This could be a feasible investigation route for the following studies, especially for animal experiments.

## Conclusions

This is the first study presenting a multicentre, retrospective case-control study on serum metabolic profiling using non-targeted UPLC-MS/MS to decipher biomarkers for ROP. We propose that creatinine, ribitol, ornithine, 10-undecenoate (11:1n1), picolinoylglycine and glutamate, gamma-methyl ester are potential biomarkers for ROP. Furthermore, creatinine and ribitol are strongly correlated with the severity of ROP. Since these metabolomic changes can be detected between the asymptomatic phase and disease phase, metabolic profiling allows stratification of ROP risk in individuals with latent ROP into high and low-risk individuals, facilitating treatment prior to development of disease pathology when the vessel proliferation is low.

Along with identifying high-risk individuals for prophylactic treatment, these risk signatures have potential value for clinical screening of new intervention measures. Selecting such individuals for participation has the potential to increase the power and benefit of clinical screening, and saving medical resources, thereby increasing treatment effectiveness.

Undoubtedly, before practical in clinical application of a metabolomic signature further studies are necessary. However, blood metabolomic profiles can provide biological information from an ongoing disease individual. We are confident that this study will provide insights into the predictive biomarkers for ROP, and theoretical rationale for the future screening and therapeutic strategies for the ROP.

## Supporting information

https://dx.doi.org/10.17632/7r7xt6pf6x

## Data Availability

All data that support the findings of this study are available within the article and Mendeley dataset. The Supplementary Information, Python code and Metabolomic Data are available in https://dx.doi.org/10.17632/7r7xt6pf6x, Doi: 10.17632/7r7xt6pf6x. Its Demographic Data File are available from the corresponding author upon reasonable request.

## Acknowledgments

The authors thank Marcus Fruttiger from Institute of Ophthalmology at University College London for advice and assistance. Also, authors thank the dedication of the Shenzhen ROP Screening Cooperative Group, which includs, but not limited to the neonatal intensive care units of the following hospitals: Shenzhen People’s Hospital, Peking University Shenzhen Hospital, Shenzhen Children’s Hospital, Shenzhen Maternal and Child Health Hospital, Shenzhen Luohu Maternal and Child Health Hospital, Shenzhen Nanshan Maternal and Child Health Hospital, Baoan Maternity & Child Healthcare Hospital of Shenzhen,Baoan People’s Hospital of Shajing Shenzhen, The People’s Hospital of Shajing Shenzhen, Huizhou Central People’s Hospital, Guangdong Provincial Maternal and Child Health Hospital, Huizhou Sixth People’s Hospital, Huizhou Third People’s Hospital, Chaonan District Mingsheng Hospital of Shantou, Shantou Second People’s Hospital, The Second Affiliated Hospital of Shantou University, Jieyang Huilai People’s Hospital, Puning Maternal and Child Health Hospital, Puning People’s Hospital, Zhanjiang Central People’s Hospital, Affiliated Hospital of Guilin Medical College, Guilin Maternal and Child Health Hospital. All of the study participants are thanked for making this research possible. Special thanks go to Dr.Haibo Peng, Dr.Chaohui Lian and nurse Sisi Luo. Special thanks also go to research assistant Panpan Sun.

Supported by Shenzhen Science and Technology Innovation Commission basic discipline layout project, China (JCYJ20170817112542555), and Medical and Health Projects of Sanming.

## Author contributions

Y.Y., J.Z., G.Z., C.L., W.W., and H.H. design the work and acquired multi-center samples. Y.Y., Q.Y. and Y.Z. performed experiments, analyzed data, and wrote the manuscript. All data that support the findings of this study are available within the article and Mendeley dataset. The Supplementary Information, Python code and Metabolomic Data are available in “https://dx.doi.org/10.17632/7r7xt6pf6x”, Doi: 10.17632/7r7xt6pf6x. Its Demographic Data File are available from the corresponding author upon reasonable request.

## Supplementary Information

**Supplementary Figure 1.**
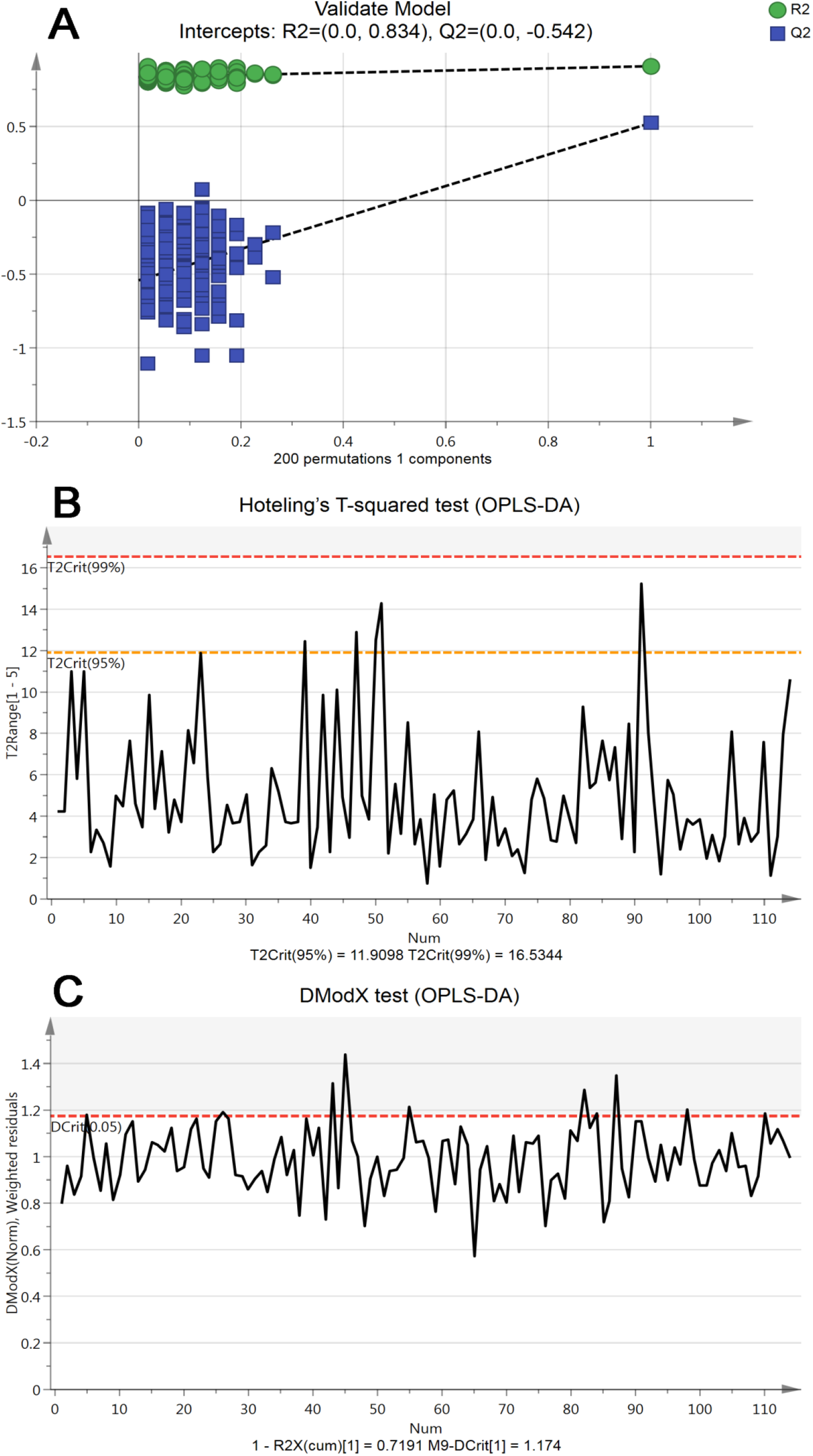
Validity tests for the OPLS-DA model. (A) Permutation analysis plotting R2 and Q2 from 200 permutation tests in the OPLS-DA model. The y-axis shows R2 and Q2, whereas the x-axis shows the correlation coefficient of permuted and observed data. The two points on the right represent the observed R2 and Q2. Cluster of points on the left represents 200 permuted R2 and Q2. Green and blue dots represent R2 and Q2 values, respectively. Dashed lines denote corresponding fitted regression lines for observed and permutated R2 and Q2. (B) Hoteling’s T-squared test revealed that most samples did not show deviation, except participants 40 (T2 = 12.43), 48 (T2 = 12.90) 53 (T2 = 14.26) and 96 (T2 = 15.23) that exceeded 95% confidence interval [CI] level but lower 99% CI level, three of these deviators were from the ROP group and one from the non-ROP group. Red and orange horizontal dashed line denotes 99% and 95% CI level, respectively. (C) DModX test plot presented that most samples did not show severe deviation in DModX, five moderate outliers being participant 43 (1.31), 45 (1.44), 55 (1.21), 82 (1.29), 87 (1.35), respectively. Red horizontal dashed line denotes 95% CI.

**Supplementary Figure 2.**
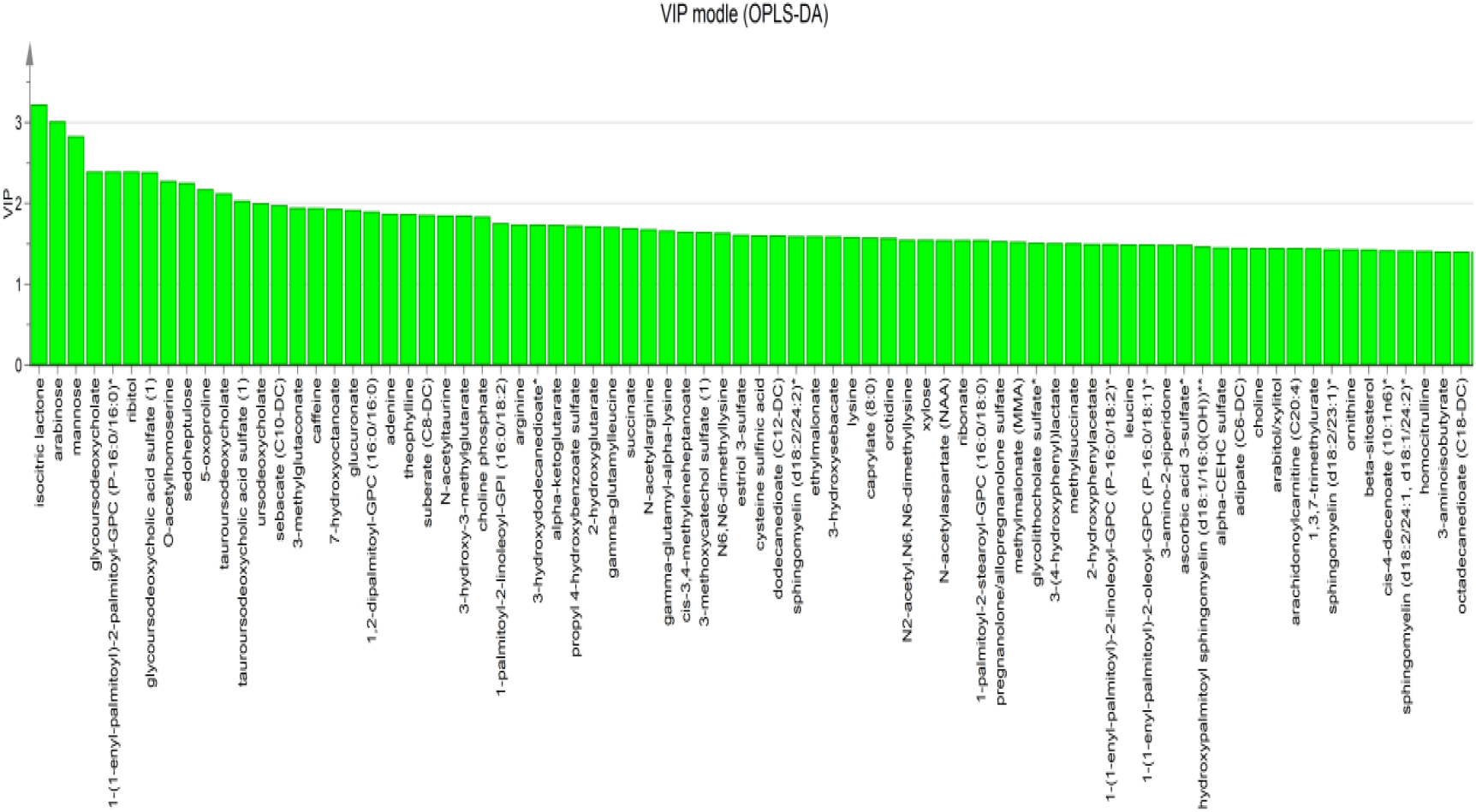
Metabolites’ contribution plot based on OPLS-DA model. Contribution plot shows the contribution of each metabolites in the previously built OPLS-DA model. According to the plot, there are 649 metabolites perceived as important with VIP value greater than 0.5 (not shown all). Only 78 metabolites were shown in the graph.

**Supplementary Figure 3.**
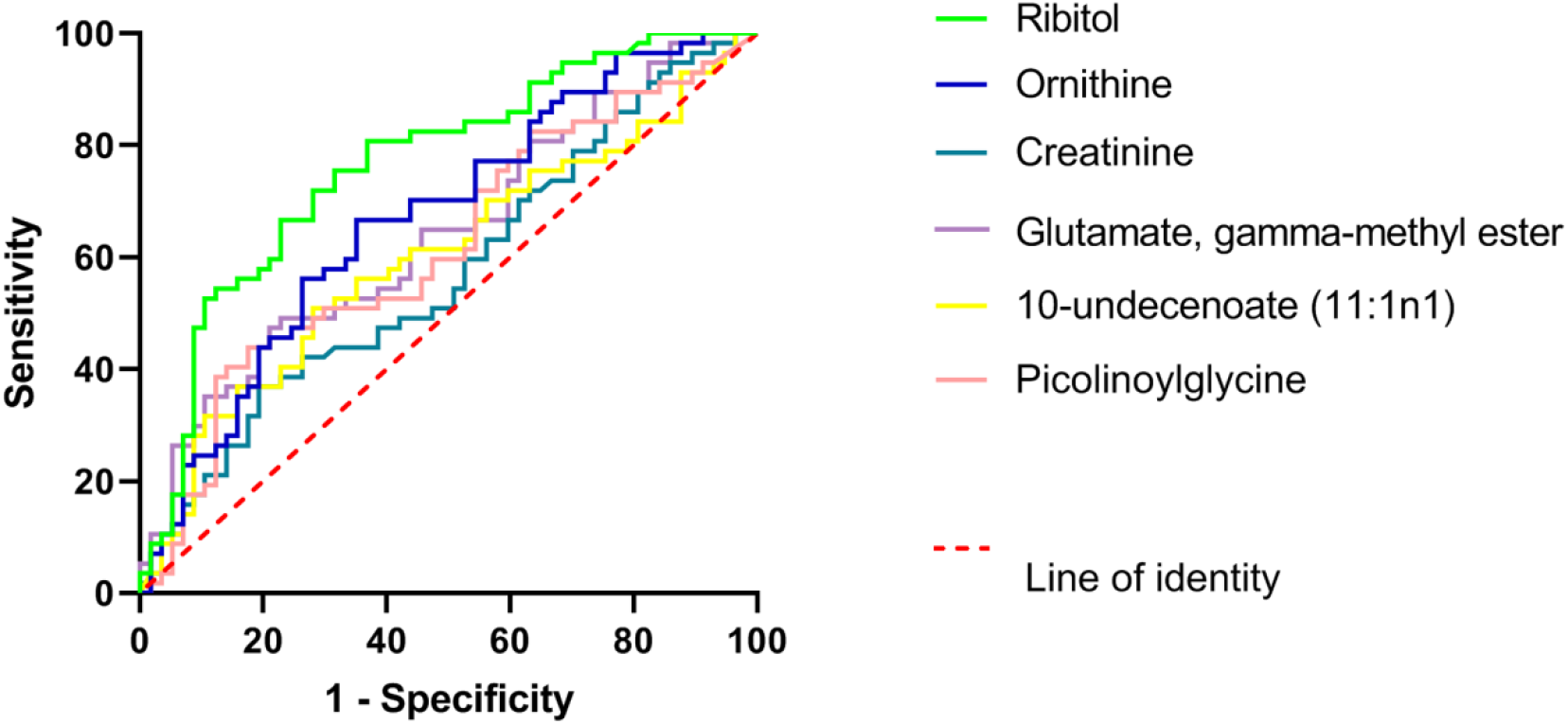
ROC of potential discriminant metabolites for ROP. Ribitol is the best among all potential discriminant metabolites at predicting ROP with high sensitivity (71.9%) and specificity (71.9%); followed by ornithine (sensitivity = 66.7% and specificity = 64.9%), which is notably inferior at predicting than ribitol.

**Supplementary Figure 4.**
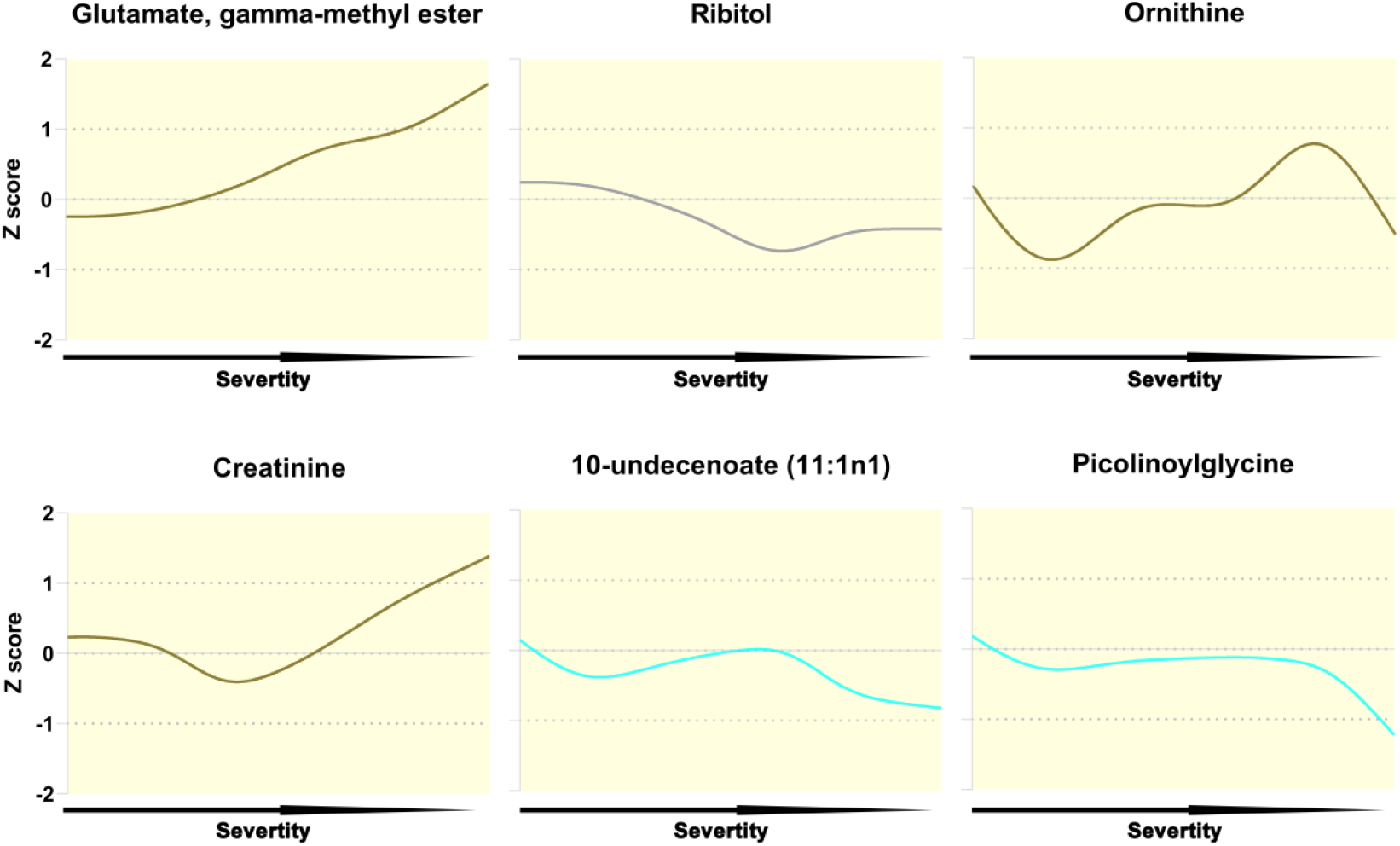
Associations of potential metabolic biomarkers with ROP severity. The associations of potential metabolic biomarkers with ROP severity are plotted with Lowess curves which colors indicate chemical classes: Amino acids = brown; carbohydrates = grey; lipids = light blue. Y-axis denotes standardized values (z-score), while the x-axis is the ROP severity. As the figure shows, the level of glutamate, gamma-methyl ester increased with the severity of the disease; ornithine and creatinine elevate with variation. However, ribitol, 10-undecenoate (11:1n1) and picolinoylglycine decreased with severity.

**Supplementary Table 1.**
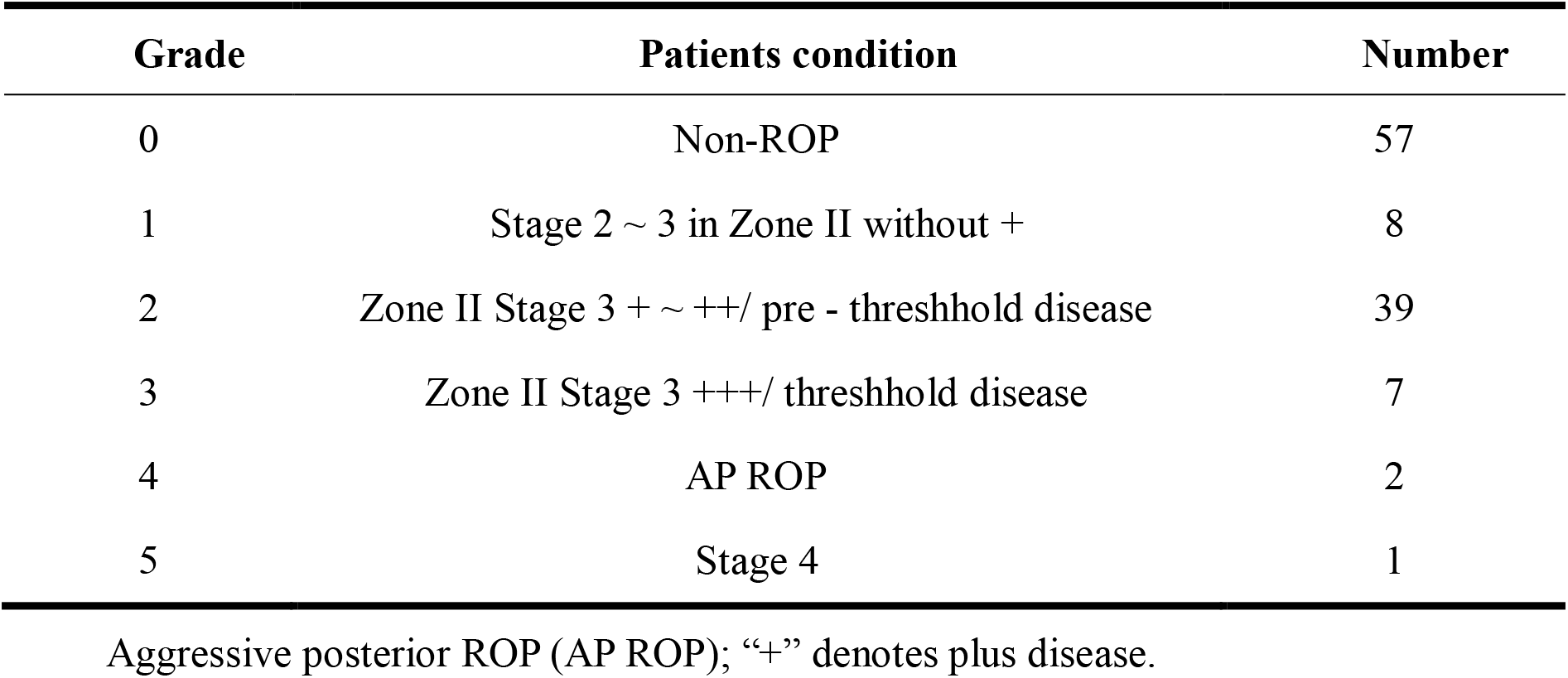
ROP severity category.

| Grade | Patients condition | Number |
| --- | --- | --- |
| 0 | Non-ROP | 57 |
| 1 | Stage 2 ~ 3 in Zone II without + | 8 |
| 2 | Zone II Stage 3 + ~ ++/ pre - threshold disease | 39 |
| 3 | Zone II Stage 3 +++/ threshold disease | 7 |
| 4 | AP ROP | 2 |
| 5 | Stage 4 | 1 |

Aggressive posterior ROP (AP ROP); “+” denotes plus disease.

**Supplementary Table 2.**
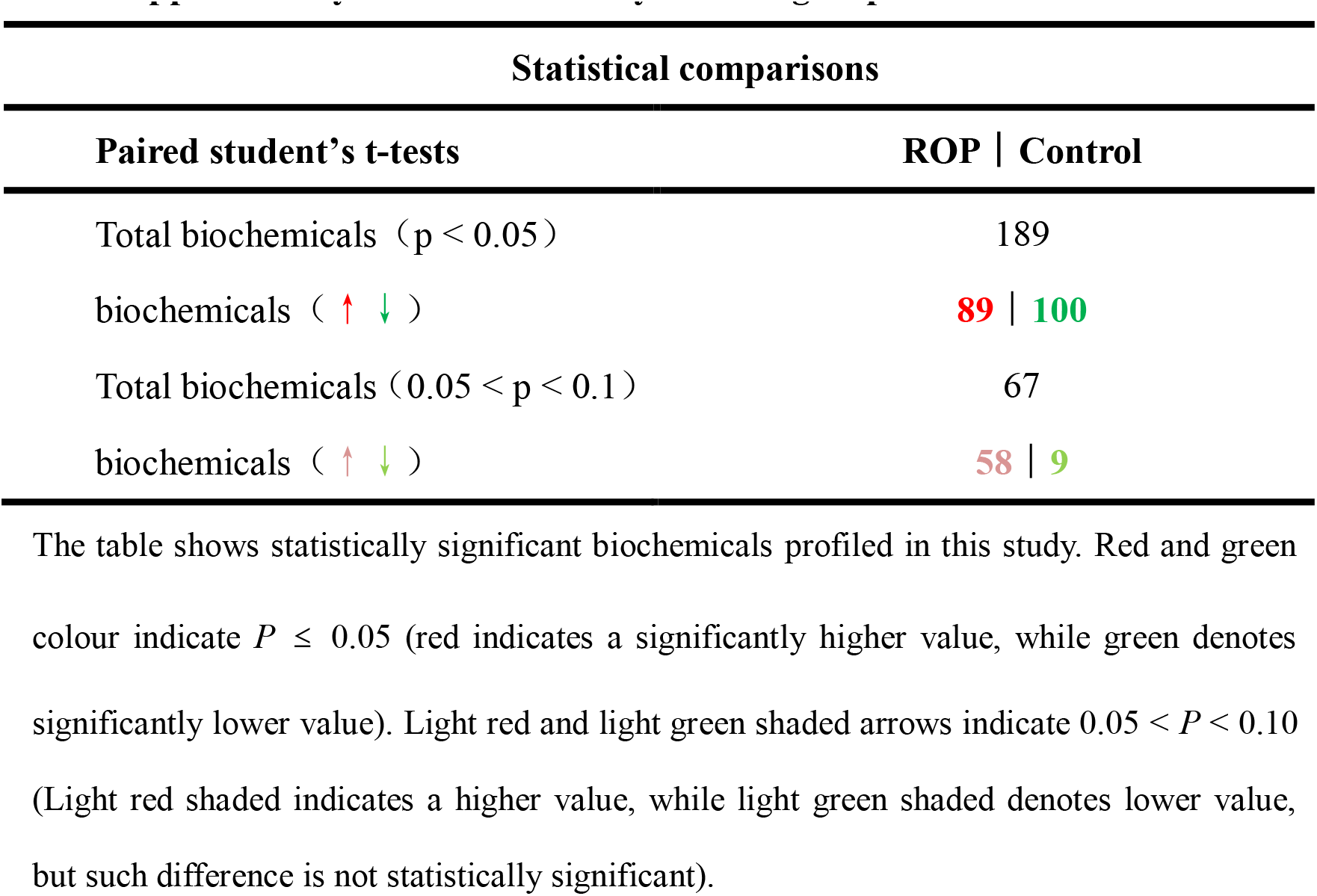
Summary of inter-group metabolomic differences.

**Supplementary Table 3.**
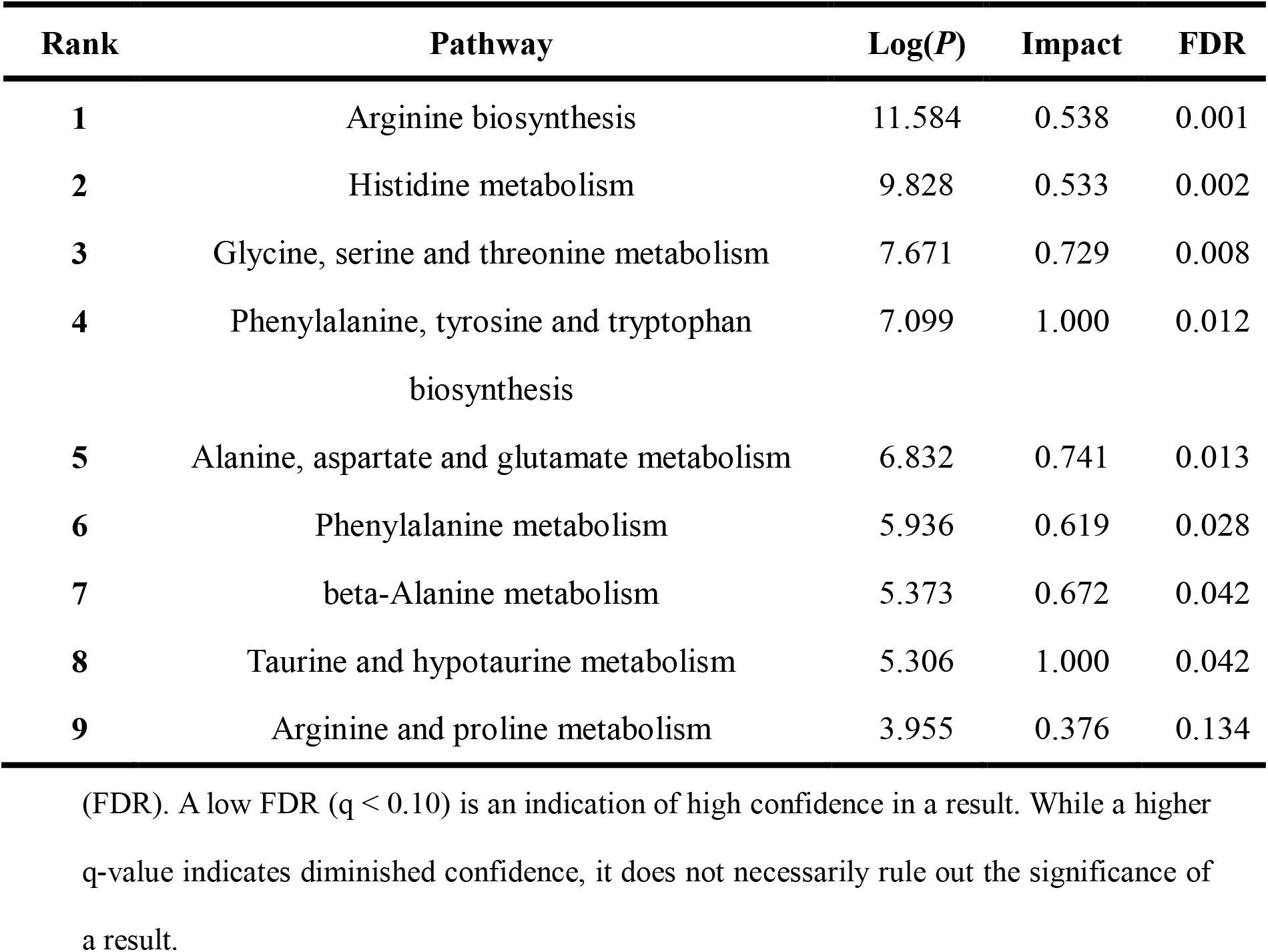
Enriched pathways. The top nine pathways are shown with -Log(*P*), impact and false discovery rate.

