## Supplementary material for "Comparative analysis unveils novel changes in serum metabolites and metabolomic networks of retinopathy of prematurity infants": https://dx.doi.org/10.17632/7r7xt6pf6x: Supplementary Informationú¿tablesú⌐.docx

**Tables**

**Supplementary Table 1. ROP severity category**

| **Grade** | **Patients condition** | **Number** |
| --- | --- | --- |
| 0 | Non-ROP | 57 |
| 1 | Stage 2 ~ 3 in Zone II without + | 8 |
| 2 | Zone II Stage 3 + ~ ++/ pre - threshhold disease | 39 |
| 3 | Zone II Stage 3 +++/ threshhold disease | 7 |
| 4 | AP ROP | 2 |
| 5 | Stage 4 | 1 |

Aggressive posterior ROP (AP ROP); “+” denotes plus disease.

**Supplementary Table 2. Summary of inter-group metabolomic differences.**

| **Statistical comparisons** | |
| --- | --- |
| **Paired student’s t-tests** | **ROP︱Control** |
| Total biochemicals（p < 0.05） | 189 |
| biochemicals（↑↓） | **89**︱**100** |
| Total biochemicals（0.05 < p < 0.1） | 67 |
| biochemicals（↑↓） | **58**︱**9** |

The table shows statistically significant biochemicals profiled in this study. Red and green colour indicate *P* ≤ 0.05 (red indicates a significantly higher value, while green denotes significantly lower value). Light red and light green shaded arrows indicate 0.05 < *P* < 0.10 (Light red shaded indicates a higher value, while light green shaded denotes lower value, but such difference is not statistically significant).

**Supplementary Table 3. Enriched pathways.**

| Rank | Pathway | -Log(*P*) | Impact | FDR |
| --- | --- | --- | --- | --- |
| 1 | Arginine biosynthesis | 11.584 | 0.538 | 0.001 |
| 2 | Histidine metabolism | 9.828 | 0.533 | 0.002 |
| 3 | Glycine, serine and threonine metabolism | 7.671 | 0.729 | 0.008 |
| 4 | Phenylalanine, tyrosine and tryptophan biosynthesis | 7.099 | 1.000 | 0.012 |
| 5 | Alanine, aspartate and glutamate metabolism | 6.832 | 0.741 | 0.013 |
| 6 | Phenylalanine metabolism | 5.936 | 0.619 | 0.028 |
| 7 | beta-Alanine metabolism | 5.373 | 0.672 | 0.042 |
| 8 | Taurine and hypotaurine metabolism | 5.306 | 1.000 | 0.042 |
| 9 | Arginine and proline metabolism | 3.955 | 0.376 | 0.134 |

The top nine pathways are shown with -Log(*P*), impact and false discovery rate (FDR). A low FDR (*q* < 0.10) is an indication of high confidence in a result. While a higher *q*-value indicates diminished confidence, it does not necessarily rule out the significance of a result.
