## Supplementary material for "Comparative analysis unveils novel changes in serum metabolites and metabolomic networks of retinopathy of prematurity infants": https://dx.doi.org/10.17632/7r7xt6pf6x: Evaluation indexes f1.docx

Best parms: {'criterion': 'gini', 'max_depth': 9, 'max_features': 'auto', 'min_samples_leaf': 3, 'min_samples_split': 2, 'n_estimators': 10}

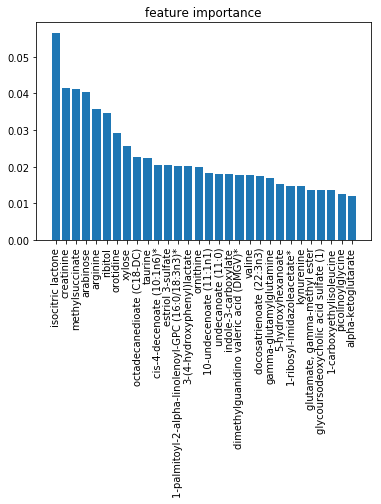

Corresponding best score: 0.8277498093058734
