## Supplementary material for "Comparative analysis unveils novel changes in serum metabolites and metabolomic networks of retinopathy of prematurity infants": https://dx.doi.org/10.17632/7r7xt6pf6x: random_forest.py

```
import pandas as pd
import config
from sklearn.model_selection import train_test_split
from sklearn.feature_selection import SelectKBest, chi2
from sklearn.ensemble import RandomForestClassifier, AdaBoostClassifier
from sklearn import metrics
import matplotlib.pyplot as plt
from sklearn.model_selection import GridSearchCV as gs


def get_content():
    """
    读取原始excel文件，将其转化为dataframe格式
    :return:
    """
    data = pd.read_excel(config.path['raw_data_path'], '5sd去除3456（已剔除）', index_col=False)
    data.to_csv('data.csv', encoding='utf-8')
    print(data.shape)
    print(data.isnull().any())  # 用来判断某列是否有缺失值
    #按列求平均值，填充
    for column in list(data.columns[data.isnull().sum() > 0]):
        mean_val = data[column].mean()
        data[column].fillna(mean_val, inplace=True)
    print("数据填充结束")
    return data

def data_process(features,labels):
    """
    对数据进行标签和特征分离，进行特征选择，划分训练集和验证集，按照二八比例划分，然后训练
    :param data:
    :return:
    """
    # 测试集大小占 20%；random_state 设置为 0 表示完全随机，默认参数下不管运行几次划分结果都是一样的（伪随机）
    features_train, features_test, labels_train, labels_test = train_test_split( features, labels, test_size=0.2, random_state=0)
    """
    特征选择
    特征选择一方面是为了降低问题的维度和模型的复杂度，另一方面是为了尽量剔除无关的变量，提高模型性能；
    从选择的数量上看，可以是按【个数】选择，也可以是按【比例】选择；本文有 11 个属性，不算多，姑且保留 9 个特征吧；
    从选择的度量方式上看，常用的指标有 卡方统计值（CHI2）和互信息（MI）；本文使用了效率较高的卡方；
    """
    sp = SelectKBest(chi2, k=9)
    features_train_selected = sp.fit_transform(features_train, labels_train)
    # 你会发现如果你在此之前还没有划分数据集，那么下面这一行可以省略，只是结果可能会稍微不同
    features_test_selected = sp.transform(features_test)
    return features_train_selected, features_test_selected, labels_train, labels_test


def traversal(features, labels):
    """
    网格搜索最佳随机森林的参数
    :param features:
    :param labels:
    :return:
    """
    params = {'n_estimators': range(10, 100,20), 'criterion': ['gini', 'entropy'],
              'min_samples_split': range(2, 5, 1), 'min_samples_leaf': range(1, 5, 1),
              'max_features': ['auto', 'log2'],'max_depth':range(1, 10, 1)}
    # scoring 选择 accuracy 值作为评估指标，cv=5 表示用 5-fold 交叉验证
    gridsearch = gs(estimator=RandomForestClassifier(), param_grid=params, scoring='f1', cv=5)
    gridsearch.fit(features, labels)
    print('\nBest parms:', gridsearch.best_params_)
    print('\nCorresponding best score:', gridsearch.best_score_)

def clf(estimator, features_train, features_test, labels_train, labels_test):
    print(features_train.shape)
    estimator.fit(features_train, labels_train)
    # 准确率
    print('Accuracy: %.3f%%' % (estimator.score(features_test, labels_test) * 100))
	# 混淆矩阵
    cm = metrics.confusion_matrix(labels_test, estimator.predict(features_test))
    print(cm)
    # 输出完整的分类结果
    print(metrics.classification_report(labels_test, estimator.predict(features_test)))
    plt.figure()
    plt.xlabel('Predicted labels')
    plt.ylabel('True labels')
    plt.imshow(cm)
    # 检测重要特征
    f, ax = plt.subplots(figsize=(7, 5))
    ax.bar(range(len(estimator.feature_importances_)), estimator.feature_importances_)
    ax.set_title("Feature Importances")
    f.show()


if __name__ == '__main__':
    data=get_content()
    features = data.drop(['组别', '配对编号'], 1)
    labels = data['组别']
    traversal(features,labels)
    features_train, features_test, labels_train, labels_test=data_process(features,labels)
    #使用最佳参数进行训练
    rf=RandomForestClassifier(n_estimators=110, criterion='entropy', min_samples_leaf= 1, min_samples_split= 5,max_features='auto',max_depth=11)
    clf(rf, features_train, features_test, labels_train, labels_test)
```
