## Supplementary figures and images for "Comparative analysis unveils novel changes in serum metabolites and metabolomic networks of retinopathy of prematurity infants"

### Supplementary Figure 1.tif

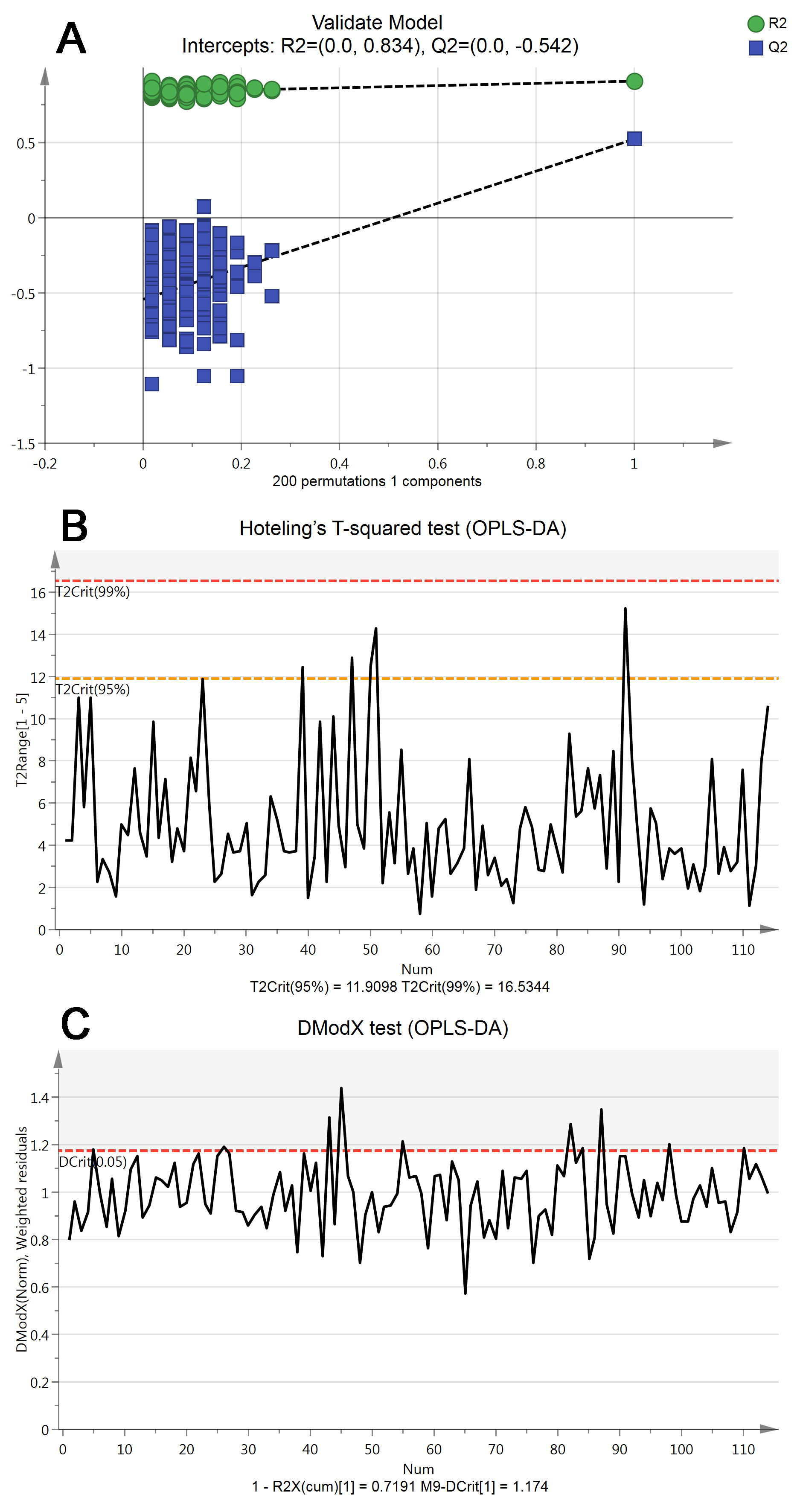

### Supplementary Figure 3.tif

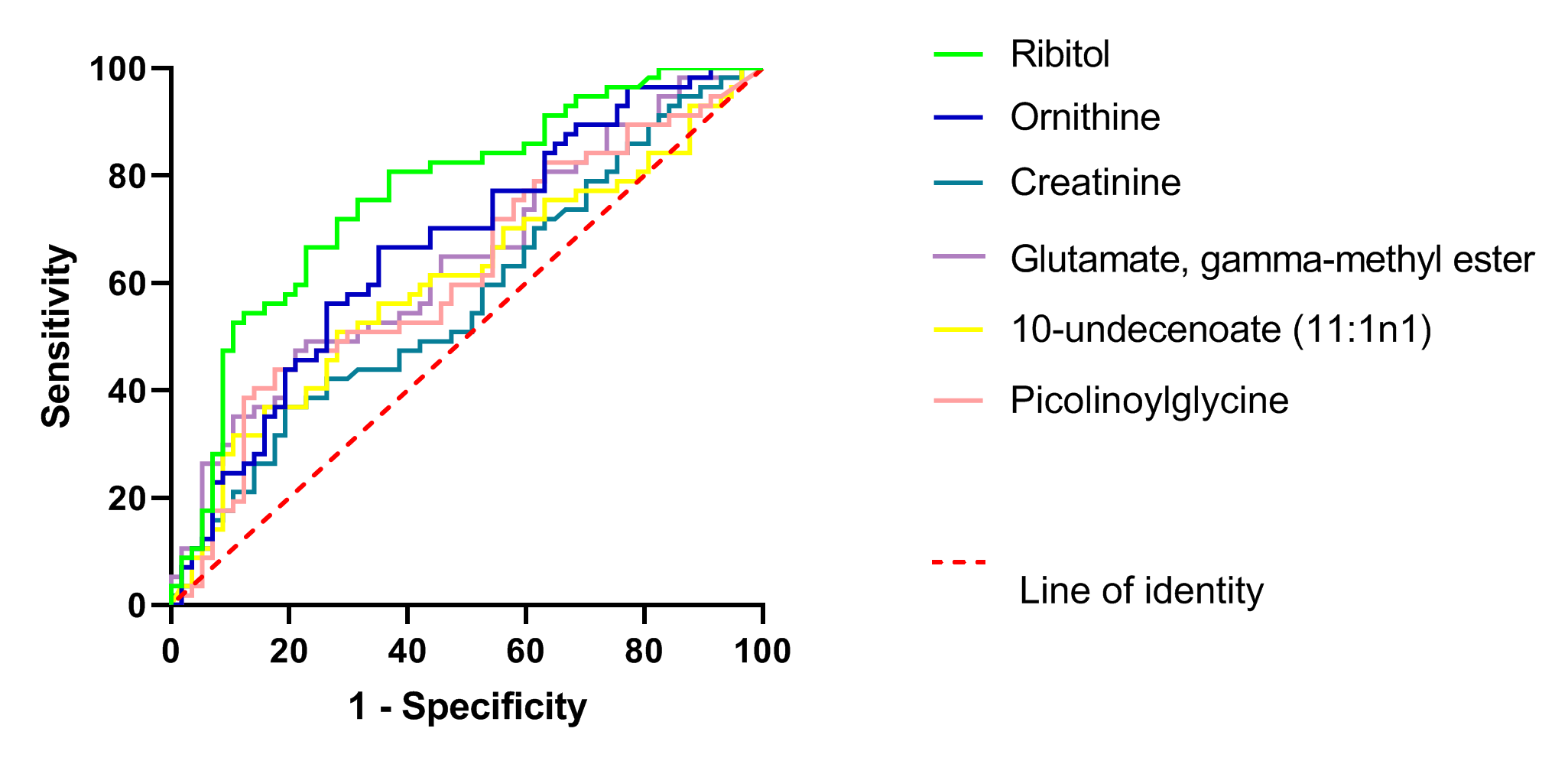
